# Giving Back: Return of Results Reports Using Digital Phenotyping Tools in Mental Health

**DOI:** 10.1101/2025.10.09.25336195

**Authors:** Manuel Soulard, Tihare Zamorano, Deven Parekh, Sara Jalali, Marie-Catherine Mongenot, Dylan Hamitouche, Ivy Guo, Chelsea Cuffaro, Katie Lavigne, Delphine Raucher-Chéné, Tosh Watson, Béatrice Schunn, Gillian Strudwick, Stefan Kloiber, Lena Palaniyappan, David Benrimoh

**Affiliations:** Department of Psychiatry, McGill University, Montreal, Quebec, Canada; Douglas Mental Health University Institute, Department of Psychiatry, McGill University, Montreal, Quebec, Canada; McGill Medical School, Montreal, Quebec, Canada; Integrated Neuroscience Program, McGill University, Montreal, Quebec, Canada; Robarts Research Institute, Western University, London, Ontario, Canada; Department of Psychology, McGill University, Montreal, Quebec, Canada; Department of Educational and Counselling Psychology, McGill University, Montreal, QC; Centre for Addiction and Mental Health and Department of Psychiatry, University of Toronto, Ontario, Canada

## Abstract

As digital phenotyping tools become more prevalent in mental health research and care, the question of how to meaningfully return data to participants, clinicians, and caregivers has grown increasingly important. Drawing from the broader DeeP-DD (Deep Phenotyping and Digitalization at the Douglas) project, this work presents the iterative development of individualized feedback reports focused on a psychiatric patient population. This paper also builds on findings from a narrative literature review to derive evidence-based practices around conveying data to participants. From the literature review, general themes such as patient engagement, visual design, ethical considerations and report content were extracted. Core design principles identified included visual simplicity, use of color, contextualized interpretation, and ethical considerations, which were compiled and helped guide the presentation of data gathered from participants in user-friendly formats. The reports created from these findings are based on both digital and clinical data, and their design was informed by key findings from the narrative review and developed through a participatory process involving patients, clinicians, and caregivers. Findings from early user feedback and existing literature suggest that well-designed reports can foster greater understanding, trust, and behavioral engagement in research and clinical care. The review also highlights important gaps, particularly in caregiver-focused communication, and discusses future directions including developing similar reports for both physicians and caregivers and the use of digital report platforms. By addressing both design and ethical considerations in a real-world setting, this work contributes to the field of return-of-results practices in psychiatric digital health.

## Introduction

In psychiatry, despite years of phenomenological and neurobiological research, there remains a lack of comprehensive and clinically actionable tools for the identification, monitoring, and staging of most mental illnesses (Clarke et al., 2024). In major depressive disorder, for example, standardized rating scales, such as the PHQ-9, are often used in measurement-based care (MBC) (Coley et al., 2020) since it has been shown to improve treatment and clinical outcomes, achieving higher rates of response and remission in randomized controlled trials compared to standard care (Zhu et al., 2021). Despite its proven efficacy, MBC is still not commonly used in routine psychiatric practice; surveys indicate that fewer than half of behavioral health providers use MBC regularly (Keepers et al., 2023). One contributing factor to this low uptake is the lack of well-designed tools, many clinicians cite poor usability, inadequate integration with electronic health record systems, insufficient training, and workflow disruptions as barriers to implementing MBC in practice (Hong et al., 2021). Also, despite their utility, these tools also have key limitations: there is no clear guidance on monitoring after remission, and their utility is more restricted in severe, persistent illnesses like schizophrenia, where self-report may be less reliable and clinician assessments are burdensome (Elliott & Fiszdon, 2014; Mote & Fulford, 2020); (Abel & Minor, 2021).

In response to these limitations, and alongside increasing access to consumer digital technologies, there is growing interest in the use of digital biomarkers, which are data points passively collected from sensors in everyday devices such as smartphones and other ‘’wearables’’ (e.g. smart watches) (Barron, 2021; Xiao et al., 2021). For instance, actigraphy technologies often found in such devices can enable the continuous assessment of sleep patterns (Rahimi-Eichi et al., 2021), which is particularly relevant in mood disorders like bipolar disorder (Dunster et al., 2021). The integration of these digital measures refers to *digital phenotyping (Torous et al., 2016)*. The broader concept of deep phenotyping includes this digital data and adds the use of other measures, such as active measures (e.g., self-report questionnaires, ecological momentary assessments), alongside clinical, historical, and sometimes biological data, like sleep patterns. This deep phenotyping approach aims to comprehensively characterize patients using reproducible metrics (Robinson, 2012).

While advances in data collection and modeling have progressed rapidly, less attention has been paid to communicating these findings to patients, caregivers, and clinicians. Yet, the manner in which digital results are conveyed may significantly shape clinical engagement, therapeutic alliance, and health outcomes (Carlier et al., 2012). Crucially, there is a lack of understanding of how best to engage patients and caregivers with the data they are providing and how to present it in order to support self-management in the clinical context. This current narrative review therefore aims to answer a key question in the field: how should results be communicated to patients, caregivers, and clinicians? At the same time, several other secondary questions are important and remain unanswered: What visual formats and timing are most effective for presenting digital phenotyping results? Does sharing data enhance patient engagement? What ethical considerations need to be addressed when communicating reports? These are essential questions for future research if deep phenotyping is to fulfill its promise as a tool for personalized, measurement-based mental health care.

‘’Returning results’’, in this study, refers to the process of communicating findings from deep phenotyping to patients, clinicians, and caregivers. It encompasses various types of results, including raw data, processed data, and actionable insights. The importance of returning results lies in its potential to inform clinical decision-making, enhance patient understanding, and promote engagement in health management (Shen et al., 2024). Beyond its clinical and relational benefits, returning results also reflects core principles of open science, an approach gaining traction in psychiatry and other fields, by increasing the transparency and accessibility of research for participants (National Academies of Sciences, Engineering, and Medicine et al., 2018).

As part of the broader DeeP-DD project (*Deep Phenotyping and Digitalization at the Douglas*), which is an ongoing initiative aimed at advancing research in the fields of digital and deep phenotyping in mental health, the current work aims to go beyond these initial inquiries and to create reports which can then be returned to participants in a useful way. Hopefully, this work will also help to assist other researchers by offering practical guidance on developing meaningful and effective patient-facing reports. We will discuss how, based on findings from the literature, digital output reports for patients were first drafted and then iteratively co-designed with feedback from clinicians, patients themselves, and caregivers to support both symptom tracking and patient self-management. We will discuss how we developed reports which synthesize complex digital data extracted from different deep phenotyping methods into actionable summaries. After further field testing and iterative design, these reports will be evaluated in a prospective clinical trial and will form the core of a broader patient, clinician and caregiver engagement strategy.

## Methodology

### Literature Review

The first section of this paper examines key themes identified through a narrative literature review on evidence-based strategies for translating raw clinical data obtained via deep phenotyping into clear, useful, and actionable reports for end users. Through this qualitative approach we synthesized the literature drawing on selected studies to provide an overview of existing knowledge, highlight recurring themes, and identify gaps in the field. English-language articles published from 2010 to the present were primarily included. Priority was given to meta-analyses, systematic reviews, and scoping reviews. Literature searches were conducted across several medical databases (Google Scholar, Embase, and PubMed) using targeted keywords such as “returning of results,” “participants,” “patients,” “providers,” “clinicians,” “caretakers,” “user experience,” “summarizing data,” and “data visualization.” 75 articles were reviewed in full, of which 40 were considered relevant based on their contributed insight into the domain of digital phenotyping or data visualization for patients, caregivers, and clinicians, both within and beyond psychiatric contexts.

### Overview of the Deep Phenotyping and Digitalization at the Douglas Project

The second section of this article outlines the process undertaken to develop the initial DeeP-DD patient reports. DeeP-DD is an ongoing feasibility study at the Douglas Mental Health University Institute in Montreal, Canada. Its aim is to explore digital phenotyping in a realistic transdiagnostic population including individuals with early psychosis, schizophrenia/schizoaffective disorder, bipolar disorder, anxiety, unipolar depression, and other conditions (e.g., personality or substance use disorders). Outpatients between 15-35 years old were recruited from the Clinical High-Risk for Psychosis, First-Episode Psychosis, Bipolar Disorder, and general Neuropsychiatry Clinics, either through clinician referral or self-referral via clinic advertisements. Participants are monitored for 6 months and up to 2 years using both digital (questionnaires, smartphone app based on the open-source Mindlamp platform (Bilden et al., 2023)) and non-digital (chart review, clinician ratings, interviews) tools to capture a broad range of observable measures. Written informed consent was obtained from all participants. Those providing actigraphy data received a GENEActiv wristband (Activinsights, UK) with instructions, and were compensated for wearing the device and completing questionnaires. The study was approved by the West-Central Montreal health authority ethics board and conducted in accordance with the Declaration of Helsinki and Tri-Council Policy Statement.

Being part of this currently ongoing study, the created reports will be used to field-test returning data in different formats, first to patients and clinicians, and then to caregivers. Their feedback will be used to inform future implementation, explore optimal sampling frequencies across diagnoses, and develop deep phenotyping practices to improve care.

### Creation of Reports

The data used to generate the reports were drawn from multiple sources. Mood and anxiety were assessed using the patient-rated PHQ-9 (Patient Health Questionnaire Nine) and GAD-7 (Generalized Anxiety Disorder Seven) questionnaires, respectively, which participants completed on REDCap every week (Harris et al., 2009; Kroenke & Spitzer, 2002; R. L. Spitzer et al., 2011). Substance use was evaluated via the DAST-10 (Drug Abuse Screening Test) and AUDIT (Alcohol Use Disorders Identification Test) questionnaires bi-annually (*AUDIT : The Alcohol Use Disorders Identification Test : Guidelines for Use in Primary Health Care*, 2001; Shirinbayan et al., 2020). Psychotic symptoms were measured using the SAPS/SANS (Scale for the Assessment of Positive Symptoms/Scale for the Assessment of Negative Symptoms) every month (Andreasen, 1984, 1989), Functional impairment was assessed with the WHODAS (World Health Organization Disability Assessment Schedule) every 3 months by participants, while global illness severity was captured through the clinician-rated CGI-S (Clinical Global Impression-Severity) monthly (Guy, 1976). The SAPS/SANS and CGI-S were completed by clinicians or an experienced research assistant on REDCap. We are actively working to replace clinician-rated measures as these are not sustainable beyond simple measures, such as the CGI, in the long run; measures such as the SAPS/SANS were collected for validation purposes. For self-rated questionnaires (such as the PHQ-9 or GAD), participants received an invitation via email through the secure REDCap platform. These questionnaires are scheduled to be sent automatically, with varying frequencies as detailed above. The timing for subsequent questionnaires was determined by the completion date and time of the baseline questionnaires.

Actigraphy data was collected using the GENEActiv actigraphy watch from Activinsights which is a compact, wrist-worn accelerometer and data logger that can also be worn on other body sites. It records limb and body movements during daily activities and sleep, storing motion data internally. It was worn by participants for one month, then returned to research assistants for data extraction which was done using the validated GENEActiv PC Software (version 3.3). This device tracked physical activity metrics (steps variability, average sedentary time, average light activity, average moderate activity and average vigorous activity) and sleep metrics (sleep variability, mean sleep time onset, sleep time onset, variability, rise time variability, mean rise time and average sleep efficiency) (Activinsights Limited, 2023). Sleep and physical activity features were extracted from the raw movement data using GENEActiv’s default R Markdown analysis tools (Selbie, 2024). Two scripts developed by lab members were used to generate plots summarizing sleep quality, physical activity levels, and symptom changes for participants, based on raw actigraphy and REDCap data (Hamitouche et al., 2025). Furthermore, more recent versions of the reports incorporate digital and social activity data, which will be gathered through the McGill-adapted version of the mindLAMP smartphone application (Bilden et al., 2023). This platform passively collects metrics such as screen and app usage, as well as indicators of social activity like phone call and text message patterns.

Study data were stored on a secure, institution-approved cloud server located in Canada, in compliance with applicable regulations. All data were de-identified at the point of collection and linked only to patient or clinician codes; no personal identifiers were included in the research files. The code key linking identifiers to research data was retained solely by the principal investigator. Access to coded data was restricted to the research team, the study sponsor and its representatives, and regulatory or institutional bodies (including the Research Ethics Board of the CIUSSS de l’Ouest-de-l’Île-de-Montréal), all of whom adhered to established privacy regulations. For safety oversight, a copy of the signed consent form was integrated into participants’ medical records, while original consent forms were securely stored in locked cabinets on site. De-identified research data may be published or presented, but no information allowing participant identification was disclosed. Data were retained for a minimum of 7 years and a maximum of 9 years following collection, in accordance with institutional policy. Participants retained the right to access their study file and request corrections, although early disclosure of certain information could necessitate withdrawal to preserve study integrity.

For the creation of the reports themselves, once guiding principles from the literature review were collected, a mock version of the patient report (see Supplementary Material) was developed to obtain preliminary feedback from the research team and Design Committee in June 2024. Once patient data began to be collected from participants enrolled in the DeeP-DD study, report creation transitioned to the web platform Canva to enhance visual quality and consistency, and a first complete version of the report was created in October 2024. Design decisions, such as the selection of symptom categories, initial layout, color schemes, and the use of line graphs and iconography, were informed by existing literature on data visualization preferences within healthcare settings (Lor et al., 2019; Midway, 2020; Sayeed et al., 2021; Turchioe et al., 2019). Other elements, however, were modified or removed based on suggestions from users and the involvement of a Design Committee, particularly in terms of improving clarity, accessibility, and perceived relevance of the content.

This Design Committee was composed of 8 clinician and/or researchers, 5 students from the Research Team, 5 people with lived-experience, including youths, and 3 caregivers of people with lived-experience.clinicians, patients, caregivers, and researchers. Members were drawn from within the Douglas Hospital ecosystem in Montreal, Quebec as well as from external centers with experience in digital phenotyping or digital integration. The Design Committee was formed through an open call to physicians and to the Center for Excellence in Youth Mental Health lived experience councils. All individuals who expressed interest were invited to join. The Committee met monthly at first to generate initial reports and then quarterly to review the progress of the project and provide feedback.

Feedback from patient-users was also collected after every report iteration through semi-structured interviews completed by research assistants and self-report Patient Experience Questionnaires administered monthly through a secure RedCap link (see Supplementary Material). For the first round of reports, five participants with different diagnoses (Bipolar affective disorder, First-Episode Psychosis and Personality Disorder) received a report, and all agreed to participate in the semi-structured interview. For the second round of reports, ten participants received a report, and four completed a second round of interviews (including two from the initial group). The study-specific Patient Experience Questionnaires assessed perceptions of various components of the project, including the utility and clarity of the reports. Of the 14 participants enrolled at the time of writing, eight completed the questionnaire at least once. Respondents had a mean age of 26.75 years and were 75% male (n = 6). Primary diagnoses included first-episode psychosis (FEP; n = 5), personality disorder (n = 1), bipolar disorder (n = 1), and clinical high risk for psychosis (CHR-P; n = 1). Results reflect pooled observations across report versions 1 to 3. Among the eight participants, one completed the experience questionnaire twice, resulting in an overall total of 9 responses.

A total of 3 versions of the reports were created at the time of writing and a total of 11 patient participants saw and provided feedback on them both through semi-structured interviews and Patient-Experience Questionnaires. In the initial iterations, based on guidance from the approving ethics board, reports were shared directly with patients by the physician in order for the information in these preliminary reports to be fully explained and to avoid any patient misunderstandings during the report refinement phase. After this initial review, subsequent reports are sent to participants via email. Our longer-term goals include refining the reports to the point where they can be provided directly to patients with no or minimal training in order to improve scalability, while balancing this against the importance of improving shared decision making (Benrimoh et al., 2021; Popescu et al., 2021; Tanguay-Sela et al., 2022). Based on feedback, a second report version was created in December 2024, keeping effective elements and adding improvements such as clearer tables, better labeling, and more detailed questionnaire data. A third version followed in February 2025, focusing on design refinements, updated graph layouts, expanded symptom tables (including psychotic symptoms), and additional substance use data. Further design details are described in the "Results - Design Principles and Report Development" section. Report creation for clinicians and caregivers is planned as part of our next steps.

## Results

The first part of our results focuses on insights from the narrative literature review, which highlighted evidence-based approaches for communicating complex psychiatric and biometric data to patients in a clear and accessible way. These findings underscored the potential of effective data visualization to improve understanding, promote self-management, and strengthen collaborative care relationships among patients, caregivers, and healthcare providers. They are organized around a series of core themes that serve as guiding principles for report creation: value of returning reports, patient engagement, visual design, ethical considerations, report content, clinical utility for physicians, and delivery medium.

The second section of our results describes the development process of our reports directly informed by the conceptual foundations and content decisions gathered from the review and user feedback.

### Value in Returning Reports

There is growing evidence that patients can derive value from access to their own health data, particularly when it is presented in a clear and intuitive manner as highlighted by (Krägeloh et al., 2015; McElfish et al., 2018). In a scoping review from (Krägeloh et al., 2015)) they examined the impact of patient-reported outcome measure (PROM) feedback in mental health care. While traditionally collected through self-report questionnaires, PROMs are increasingly complemented or even supplanted by digital phenotyping approaches (Raballo, 2018). This integration enables a more dynamic, real-time understanding of patient outcomes and symptom trajectories (Insel, 2017). Unlike previous reviews reporting limited effects (Boyce & Browne, 2013), (Krägeloh et al., 2015). found that establishing a formalized process increases the likelihood that feedback will be reviewed with patients, additional elements of procedural structure may also play a role, including the availability of computerized support tools, the frequency with which feedback is provided, and the extent to which PROM results are discussed among clinicians. It highlights that returning results to users in a structured manner can enhance patient-centered care by promoting active participation and improving communication.

(Sayeed et al., 2021)) aimed to characterize participant preferences and expectations regarding the return of individual research results within the Project Baseline Health Study, a longitudinal, multicenter cohort study enrolling participants from the general population and groups at elevated risk for heart disease and cancer. Relevant outcomes included the finding that 54.9% of participants reported being "excited to learn more" about their health, and 36.4% described themselves as "curious" when presented with the prospect of receiving individual results, with primary motivations including learning new information (72.5%), improving health (72.0%), and identifying disease risk (70.1%). These findings highlight the motivation of some patients to become active participants in their own care, a cornerstone of patient-centered approaches in mental health and in alignment with core pillars of recovery-oriented mental health care, such as empowerment, personal responsibility, and autonomy. Frameworks like CHIME highlight how such engagement fosters meaningful participation and improved outcomes (Leamy et al., 2011). However, Sayeed et al. (2021) also reveal important nuances in patient preferences. While the majority expressed enthusiasm, a smaller subset reported feelings of nervousness or distress when confronted with complex or ambiguous results (7.4%), sharing concerns about the unintended consequences and logistical burden of returning results. Sayeed et al. (2021) also emphasized that not all patients want access to the full spectrum of their data, some preferring only actionable or contextualized findings rather than raw metrics. This variability aligns with (Krägeloh et al., 2015)’s findings on patient-reported outcome measures (PROMs), which suggested that unfiltered data can sometimes increase anxiety or lead to misinterpretation without adequate framing and support. Such evidence supports the need for flexibility in report design and for offering patients choices about the depth and detail of the information they receive.

### Visualization Design

#### Simplicity and Focus

We identified a series of core principles to guide effective data visualization when presenting patient-facing results (Midway, 2020). These principles emphasize clarity, accuracy, and interpretability and provide a framework for designing patient-facing visuals in different settings. The author included specific titles for his principles, shown here in quotation marks. First, prioritizing conceptual design before execution (called “Diagram First” by Midway) ensures that visualizations convey the intended core message rather than being shaped solely by technical constraints. Selecting appropriate software (“Use the Right Software”) is also critical, as advanced or specialized tools may be required to achieve the desired technical precision and aesthetic clarity. They also discussed that optimizing geometric representation (“Use an Effective Geometry and Show Data”) is essential to accurately reflect the underlying data and to enhance transparency. Pairing simple visuals with detailed captions (“Simple Visuals, Detailed Captions”) further contributes to accurate interpretation and independent understanding. Finally, they argued that iterative feedback from patients and clinicians (“Get an Opinion”) is indispensable in refining visual elements to maximize clarity and acceptability.

Literature regarding visualization design overall emphasizes principles of visual simplicity and focus, advocating for figures that communicate a single, clear message that grabs users’ attention while minimizing clutter. Other articles from the narrative review support this, indicating that line graphs and bar graphs are widely understood by both patients and clinicians (Turchioe et al., 2019); Lor et al., 2018; Hartzler et al., 2015). By emphasizing clear trajectories rather than static snapshots, these formats help patients identify meaningful trends in their health status.

#### Engaging Formats

In addition to being easier for users to understand, the literature review emphasized that incorporating visual elements such as icons and simplified graphics can further enhance engagement by facilitating interpretation, particularly in domains such as sleep, mood, and activity (Lor et al., 2018). In their study they aimed to systematically synthesize the literature on information related to symptoms visualization. Notably, (Schroeder et al., 2017) reported that displaying patient nutrient and symptom data using line graphs and bar charts prompted both patients and providers to inquire about the data collection process and to request comparative population-level information, thereby helping to foster greater interest in and engagement with care.

As described above, the consistent preference for line graphs reported across multiple studies underscores patients’ need for clarity and intuitive design. This is further reinforced by evidence showing that patients are more engaged and more likely to act on information when it is presented in simple, visually appealing formats (Chishtie et al., 2022). In their study, they highlighted that displaying biometric data over time enables users to identify patterns, such as increases, decreases, or fluctuations, thereby supporting progress monitoring and the early detection of deviations from expected ranges. This enhanced ability to track and interpret data ultimately helps empower individuals to actively participate in treatment planning, self-management, and informed clinical decision-making.

Therefore, graphics and other visual elements constitute essential design components to consider in report development, as they have a direct impact on user comprehension and engagement.

#### Strategic Color Use

Color is another critical design element. The reviewed literature highlights the effectiveness of "traffic light" color schemes to draw attention to important changes or thresholds (Turchioe et al., 2019). While vibrant palettes can increase memorability and emotional resonance, evidence also cautions that colors must be chosen deliberately to avoid misinterpretation, particularly in populations with color blindness. Furthermore, it was also one of Midway’s core principles ; the deliberate use of color (“Colors Always Mean Something”) can further reinforce meaning, improve memorability, and support accessibility, especially when using traffic light schemes or colorblind-friendly palettes. Maintaining a high data-to-ink ratio, meaning maximizing the proportion of visual elements that represent actual data, also helps avoid unnecessary embellishments and focuses the viewer on key information (Midway, 2021). Another way to integrate color in reports is through ‘’alerts’’. (Abudiyab & Alanazi, 2022; Chishtie et al., 2022) mentioned that visualized alerts highlighting significant changes or abnormalities in biometric data with visual cues, such as color-coded indicators or warning symbols, can effectively draw the user’s attention to important events. These findings suggest that color selection is not merely a subjective or aesthetic consideration but has a direct impact on user experience and comprehension. They underscore the importance of careful, evidence-informed color choices and highlight the value of incorporating user preferences into visual design decisions when developing reports.

#### Mental Health

In recent years, authors such as Turchioe, Chang, and Polhemus have made notable contributions toward addressing the significant gap in knowledge about visualization practices specifically in mental health care. Their work underscores the benefits of digital phenotyping and thoughtful visual design, offering valuable guidance for understanding and developing effective patient-facing documents.

Notably, (Chang et al., 2023), conducted a qualitative project examining how patients and clinicians experienced the integration of personal data visualizations into mental health treatment. Their aim was to explore whether and how these visual displays, derived from digital phenotyping data, could meaningfully inform care. They used semi-structured interviews with 10 patients diagnosed with depression and anxiety at a digital mental health clinic. The authors analyzed narratives through inductive thematic analysis to identify key factors that shaped users’ comprehension and utility of the visualizations. Results supported previous literature in the medical population, revealing that patients valued simplicity and focus, expressing that straightforward bar graphs and longitudinal symptom charts helped them quickly identify patterns without becoming overwhelmed by information. The study also underscored patient engagement, as many participants reported that seeing their own data in clear visual form motivated them to reflect on mood fluctuations, initiate conversations with clinicians, and feel validated in their progress. Patients highlighted that use of color, for example, color-coded segments denoting varying levels of activity or mood, were helpful because these cues immediately signaled areas needing attention and made complex temporal data easier to interpret. (Polhemus et al., 2022) echo these findings. In their systematic review of 31 studies examining how people with chronic neurological and mental health conditions experience data visualizations from remote measurement technologies, they found that clear and simple visuals were crucial as users preferred simpler graphs that highlighted key patterns without overwhelming detail. Visuals also promoted engagement, with many reporting that seeing their own trends increased motivation, a sense of control, and validation of their experiences. The study noted again the importance of color-coded elements to help users quickly interpret changes or thresholds, making data more actionable and meaningful in managing their health.

Chang et al. (2023) further emphasized that visualizations integrated into mental health care encouraged reflection and dialogue, enhancing the therapeutic encounter by providing a shared, interpretable reference point for patients and clinicians. These findings indicate that well-designed patient-facing visuals can enhance understanding, promote collaboration, and maintain engagement in both general and mental health care, though evidence remains limited and further research is needed.

### Clinicians and Caregiver Perspectives

#### Clinician Experience

While the focus of this review is predominantly on patient perspectives, our findings also indicated that many of the themes of visual elements, engagement and report content considered beneficial for returning results to patients were also viewed as significant and actionable by physicians.

In a human-centered design study, (Hartzler et al., 2015) engaged healthcare professionals in the iterative development of a Patient-Reported Outcomes Dashboard to support improvement in post-surgical care. Clinicians emphasized the need for clear, concise visualizations that enable quick identification of trends, outliers, and comparisons to population benchmarks. Interestingly, similar to patients, they were shown to prefer bar charts and line graphs formats over pie charts and box plots. Physicians seemed to particularly prefer models with simple, static dashboards suitable for time-constrained clinical settings. More specific to clinical practice, clinicians valued the ability to filter data by patient characteristics and requested timelines to visualize individual progress. Furthermore, the study underscored the importance of also incorporating user-centered design principles and iterative feedback when developing data visualization approaches for healthcare applications used by physicians. Similarly, (Abudiyab & Alanazi, 2022) emphasized the importance that effective visualization techniques rely on interactive dashboards and clear graphical representations, including real-time displays of patient status indicators (e.g., oxygen saturation and vital signs), especially in a high or intensive care setting where parameters change quickly. Standardized color schemes to highlight deviations from expected ranges were also used.

Complementing these findings, (Chang et al., 2023) in their qualitative project also interviewed 5 mental health clinicians from a digital mental health clinic to gain insight into their experience with data visualization. They found that clinicians valued the use of color, describing how color-coded indicators quickly highlighted areas of concern or improvement, allowing them to prioritize discussion points and guide treatment decisions efficiently. Overall, the study illustrates how well-designed visualizations can support clinicians in delivering more effective, patient-centered care. Clinicians also emphasized that simplicity and focus in visuals, such as straightforward bar graphs and symptom timelines, helped shared-decision making as it made it easier to explain trends during sessions and reduced time spent interpreting raw data. Clinicians also noted that these visuals strengthened engagement, as patients were more likely to ask questions, reflect on their progress, and participate actively when shown clear graphical feedback.

Although the literature on visual design in mental health settings remains sparse, existing findings suggest that color schemes and graph-based formats can enhance comprehension. Mental health care, however, often involves more subjective, longitudinal, and symptom-based data than many general medical fields, which may require different approaches to nuance and context. Even so, the parallels in feedback from mental health clinicians and general physicians indicate that core design principles, such as clarity, intuitive visuals, and well-structured information, are broadly applicable and can still guide the development of effective patient-facing reports that support understanding, interpretation, and decision-making.

#### Caregiver’s Experience

Despite the increasing involvement of caregivers in mental health support and care, our review, while not systematic, found little guidance in the existing literature on how best to return individualized health data to this group, as highlighted by (Oakley-Girvan et al., 2023). While not specifically focusing on detailed report-return, some qualitative studies (Dalstrom et al., 2025); (O’Neill et al., 2023) do suggest that caregivers value access to data, communication, and being informed; pointing towards the potential benefits of returning reports. By integrating caregivers in patient management and systematically providing them with clear, actionable summaries of patient data, such as symptom trends or progress indicators, it may enhance their sense of partnership, reduce uncertainty, and support more coordinated care. This underscores a critical area for future investigation, namely how the process of returning individualized reports to caregivers is best implemented and to what extent it is perceived as useful in supporting clinical decision-making, enhancing relational dynamics, and improving outcomes for both patients and their support network. Importantly, this process also raises ethical considerations around consent, particularly in determining when and how patients agree to share personal health information with caregivers.

### Ethical Considerations

Considering the sensitivity of the data provided by patients, ethical issues can arise when returning results. For example, the large volume of passively collected data can reveal sensitive or stigmatized behaviors, potentially affecting participants’ well-being; many findings lack clear clinical validity or utility, and may be influenced by biases in models created when systems are trained on data that do not adequately reflect the relevant characteristics of the participants in the study. It is widely acknowledged that non-representative training data can lead to biased outcomes in medical AI (Vokinger et al., 2021). Additionally, the risk of unintentionally capturing data from third parties raises serious privacy concerns (for example, as described by Shen et al (2024), Bluetooth proximity data could inadvertently identify nearby individuals, whose GPS traces might then be linked to locations associated with illicit drug use). These issues underscore the need for careful, ethical approaches to result disclosure. (Shen et al., 2024) tried to provide guidance for how digital phenotyping results should be returned in real-world settings, specific to the psychiatric field. This group began its work by considering existing ethical and legal guidance and drew from recommendations on the return of research results more broadly, with a particular focus on the return of genetic research findings. They proposed a tiered framework for returning research results, emphasizing the need to consider both clinical utility and potential harm. They recommend that results with clear meaning and low risk should generally be returned, while those with ambiguous significance or higher risk warrant a more context-sensitive approach based on participant preferences and study design. When both benefit and harm are unknown, results should be withheld until further clarity is achieved. Importantly, decisions should be made on a study-by-study basis, and participants should ideally be given choice and control over what data they receive and when. This article emphasizes the critical importance of determining which specific information should be returned to patients while underscoring that not all collected data warrants disclosure, and that decisions about which results to include in reports requires careful evaluation.

### Design Principles and Report Development

As outlined in the methodology section, the development of the DeeP-DD Patient Reports was based on the core principles for effectively communicating clinical data to patients highlighted in our literature review and summarized in Table 1. As presented in the Results above, the literature findings were important as a starting point to report creation since effective visualization of data can help users identify patterns, trends, and anomalies more easily, leading to better-informed decision-making and improved patient outcomes (McElfish et al., 2018). We were also encouraged by supporting findings that this endeavour seemed worthwhile as patients in previous work demonstrated an interest in receiving and utilizing their personal health data to inform self-care decisions and monitor health trends (Chang et al., 2023; Polhemus et al., 2022; Sayeed et al., 2021). Building on this first literature-inspired version of our report, the subsequent iterations, V2 and V3, were further refined based on the feedback collected from both our Design Committee and participating patients.

**Table 1.** Summary of Core Principles for Returning Results in Patient-Facing Reports.

| Themes | Core Principle | Description | Key Source(s) |
| --- | --- | --- | --- |
| **Patient Engagement** | Value of Access | Patients express strong interest in receiving and using personal health data to guide self-care and track health trends. | Sayeed et al., 2021 |
|  | Diverse Preferences | While most patients are excited or curious about receiving results, some report nervousness or distress when content is complex or unclear. | Sayeed et al., 2021 |
| **Visual Design** | Simplicity and Focus | Visuals should emphasize a single key message, minimize clutter, and use line graphs for trending data. | Midway, 2020 |
|  | Engaging Formats | Line and bar graphs are widely rated as the most understandable by both patients and clinicians. Icons support easier interpretation of domains. | Lor et al., 2018; Hartzler et al., 2015 |
|  | Strategic Color Use | Colors, especially traffic light schemes, can effectively highlight important changes. A high data-to-ink ratio supports clarity. | Midway, 2020; Turchioe et al., 2019 |
| **Ethical Considerations** | Tiered Disclosure Framework for Data | Importance in need to consider both clinical utility and potential harm. Results with clear, low-risk meaning should be returned by default; ambiguous or higher-risk findings should be returned based on patient preference and context. | Shen et al., 2024 |
|  | Participatory Approach | Iterative design involving patient and clinician feedback ensures relevance, acceptability, and ethical integrity. | Midway, 2020; Shen et al., 2024 |
| **Report Content** | Actionability | Patients value contextualized results that include explanations and suggestions to support behavioral change. | Sayeed et al., 2021 |
|  | Threshold-Based Alerts | Visual cues (e.g., color-coded warnings) based on validated clinical thresholds help identify and respond to symptom changes. | (Abudiyab & Alanazi, 2022; Chishtie et al., 2022) |
| **Clinician's Experience** | Time-Sensitive Design | Clinicians prefer simple, quick-to-read summaries with benchmarks and timelines that support fast interpretation with filtering and personalization options | Hartzler et al., 2015; Khairat et al., 2018; (Chang et al., 2023) |
|  | Trend and Progress Monitoring | Used trend lines to detect clinically significant variations | Hartzler et al., 2015; Abudiyab et al., 2022 |
|  | Shared Decision-Making | Visual reports should support collaborative discussion between clinicians and patients. | Hartzler et al., 2015; (Chang et al., 2023; Polhemus et al., 2022) |

Our goal was to create a concise report that focuses on clarity, ease of use, actionability and safety through the use of visual elements. Version 1 of the reports (V1) therefore consisted of a single double-sided page, divided into three sections: (1) ‘’Call to Action’’ ; monthly actigraphy measures representing behaviors more directly under patients’ control (e.g. hours of sleep, physical activity), (2) ‘’Outcomes’’ ; measures from questionnaires (e.g. mood, stress, general functioning), and (3) ‘’Additional Data’’ ; here, actigraphy results since the beginning of the study, used to provide a broader temporal overview of symptom trend. Finally, sections for digital and social activity were included in early versions of the reports, with data collection planned via the McGill-based version of the mindLAMP smartphone application; due to deployment timelines these were only utilized for patients in the later report versions, once the app had launched. Visualizations for GPS displacement data are also currently under consideration. The choice of these starting data points was informed in part by Shen’s ethical framework of returning results (Shen et al., 2024), discussions with the clinicians involved in the project, and evidence about digital phenotyping results most correlated with patient symptoms (such as social activity) (Henson et al., 2020). We chose to prioritize low-risk, more easily interpretable measures such as sleep, activity, and mood trends, while intentionally excluding more sensitive or potentially stigmatizing information like suicidal ideation or clinician ratings of participant insight. These decisions reflect a careful consideration of both clinical relevance and the ethical implications of returning complex or ambiguous data, especially in psychiatric contexts where digital phenotyping can raise concerns about misinterpretation or harm. Multiple studies in the literature also supported our decision to establish a Design Committee and to collect feedback from users throughout the development process (Midway, 2020; Polhemus et al., 2022; Sayeed et al., 2021; Shen et al., 2024). This participatory approach helped ensure that the data included in reports aligned with what participants found meaningful and acceptable to receive.

For the visual elements, Midway’s key visualization concepts directly informed the design of the initial mock report in our project (all versions of the reports can be found in the Annex). Refer to Figure 1 for an example of the current version of the report (V3). To align with elicited themes of simplicity, focus and engagement from the literature, the design team opted for clean visual layouts emphasizing line graphs for trended data, such as sleep, steps (physical activity), and psychiatric symptom trajectories (depression, anxiety and negative/positive symptoms of psychosis), as these formats support longitudinal interpretability while avoiding clutter (Hartzler et al., 2015; Lor et al., 2019; Midway, 2020). To follow strategic color use, bold color palettes were first chosen and then workshopped in partnership with the Design Committee and through participant feedback sessions, ensuring emotional resonance, accessibility, and differentiation across categories. To follow the principle of threshold-based alerts, outcome measures graphs were color-coded following a traffic light scheme and using color-blind friendly tones as a way to facilitate interpretation of symptoms severity (Midway, 2020; Turchioe et al., 2019); (Abudiyab & Alanazi, 2022; Chishtie et al., 2022). Icons were introduced to clearly mark categories and short labels explaining the categories were prioritized over heavy text to keep visual density low and viewer attention high (Midway, 2020).

**Figure 1.**
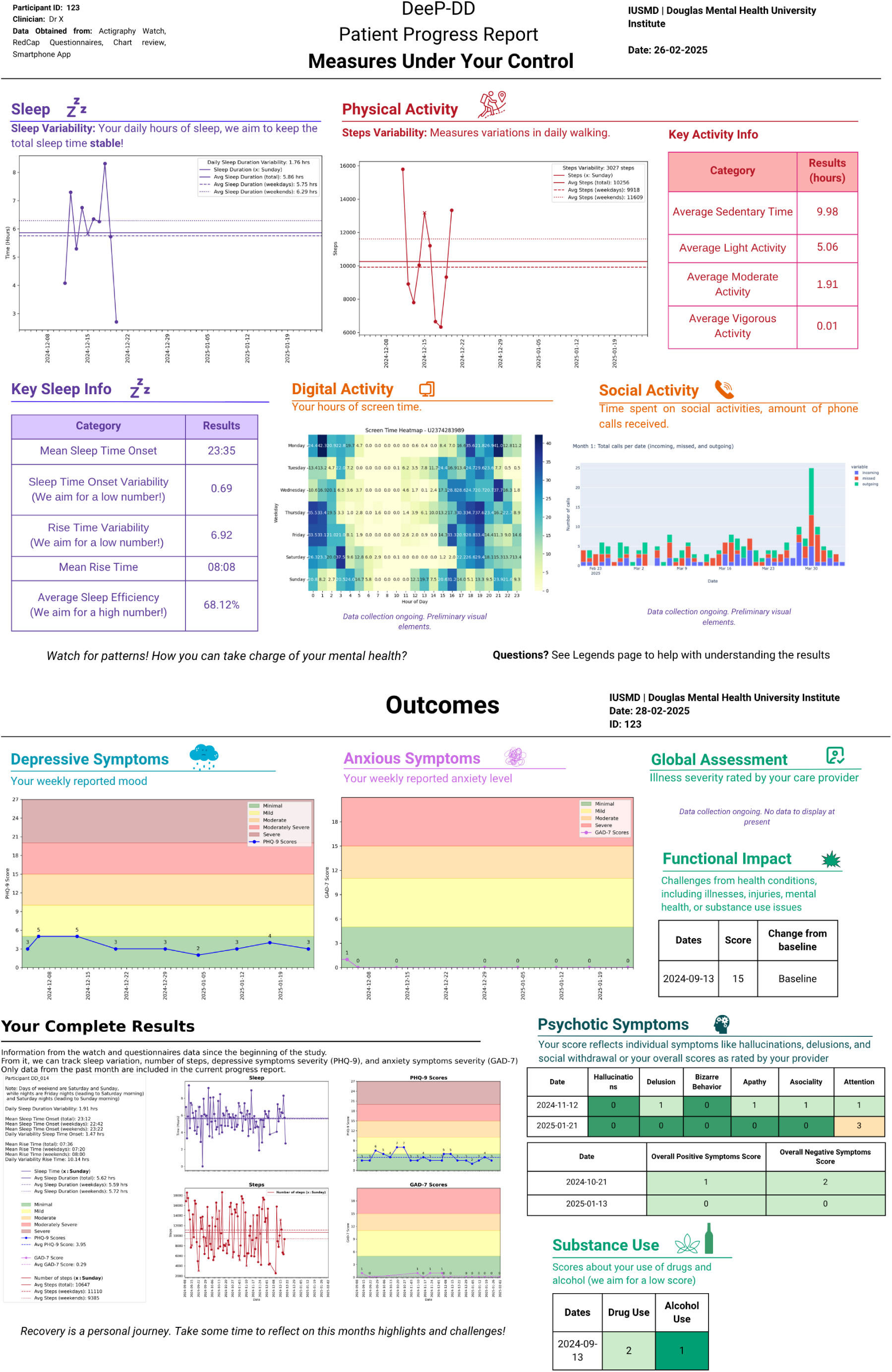
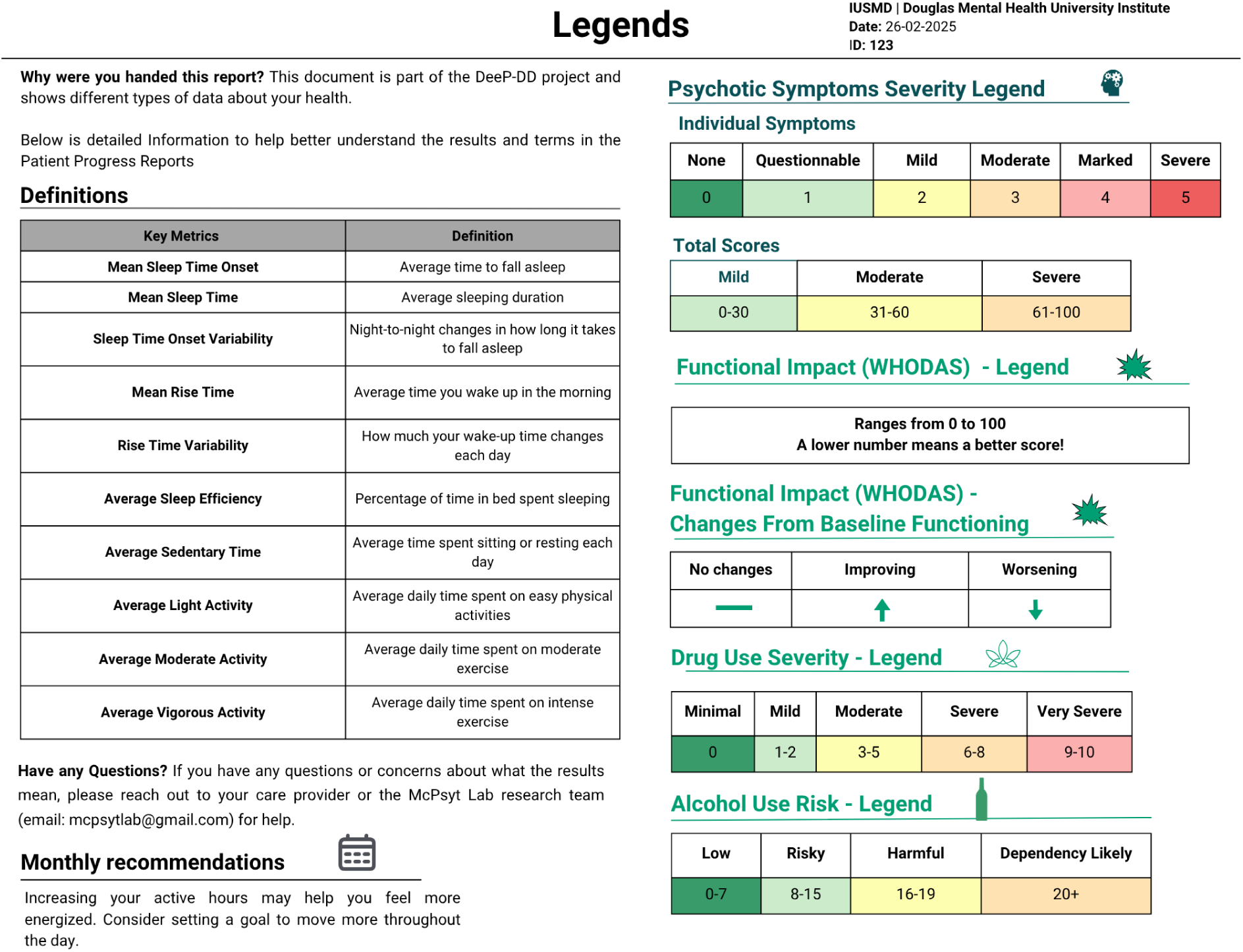
Report Version 3. **Version 3 Report Description**: Report content division revised and distributed across three single-sided pages to improve readability. First page: (1) “Call to Action” renamed “Measures under your control”. Preliminary visual elements for “Digital” and “Social” Activities were available but not finalized at the time of writing. Second page: (2) “Outcomes” and (3) “Additional data”, renamed “Your complete results” – bottom left – and now comprising self-reported scores alongside actigraphy data from study beginning. Sundays are marked with an “x” on line graphs to highlight differences between weekends and weekdays. Graphs display mean values for weekdays, weekends, and overall totals, allowing clearer interpretation of patterns across time. Both self-reported/clinician-rated score data tables and line graph keeping with the traffic light color scheme. Third page: additional section (4) “Legends” with severity scale legends and definitions of key metrics. Monthly recommendation section also moved to the last page bottom left. Each category of data still includes an icon and a short label written in a distinct color. Report’s colors reviewed to ensure consistency between each symptom category, particularly in the “Your Complete Results” section. All graphs standardized to use the same X-axis.

Each report also included one tailored monthly recommendation written by the research team, designed to translate the presented data into feasible behavioural changes, thereby further fostering patient engagement (Sayeed et al., 2021; Shen et al., 2024). The decision to emphasize only one recommendation per report aligns with the principle of simplicity and focus (Midway, 2020).

The creation process deliberately combined basic and more advanced visualization tools to balance usability and customization. Initial prototypes were first sketched using accessible tools (e.g., Excel) to quickly iterate on structure, but updated versions (V1 to V3) were generated using more advanced platforms (Canva) which was more suited to visually appealing report creation as it allowed for easy fine-tuning of layout, interactivity, and color calibration (Midway, 2020).

Moreover, the decision to maintain multiple rounds of feedback from the Design Committee as well as patients aligned with the review’s focus on adjusting design through user feedback (Midway, 2020; Shen et al., 2024). This approach helped identify user preferences that were mostly in line with the literature. For example, a preference emerged for clearer line graphs, distinct color schemes, and the use of intuitive icons to separate domains (e.g., mood, sleep, activity). The first report (V1) prompted feedback primarily on information layout and visual design. Design Committee members shared that some graphs were difficult to follow and that some parameters (e.g. sleep and physical activity) were difficult to read. They also highlighted the fact that some specific terminology used (e.g. the names of the questionnaires, sleep efficiency) was difficult to understand.

In response, from the second iteration (V2) onward, a new section was added on a second page to provide legends for severity scales for different sets of data and to provide definitions of key metrics. Some data were also converted from line graphs to tables, as this format was considered easier to read especially for the more detailed sleep and physical activity parameters (Average Sedentary Time, Mean Sleep Time Onset, etc). The table format was also used to track data that required too many lines to be put in a graph while remaining clear (various psychotic symptoms, and less frequently collected functional impact and substance use data). These tables still followed the traffic light color scheme to help track symptom severity as this feature was found helpful by users. Interestingly, although the use of tables was not specificaly included in the recommendations from the literature, this change was based on received feedback and aimed to improve clarity and interpretability for these specific sets of data.

Feedback on V2 focused on improving visual organization and clarity. Suggestions included limiting the number of colors used, assigning consistent colors to each symptom category, particularly in the “Your Complete Results” section which displayed results since the start of the study, and adding borders around graphs to better differentiate metrics such as anxiety and sleep. Notably, some feedback from the Design Committee diverged from the literature, which often recommends limiting content to a single page to enhance clarity. While the literature emphasized brevity to help reduce cognitive load, Committee members found the first page of V2 overly dense, particularly due to the addition of new tables, and expressed concern that the amount of information compromised readability. In response, content was redistributed across multiple pages to improve visual flow and comprehension. This decision reflects a deliberate trade-off: prioritizing the inclusion of more detailed, clinically relevant information over strict adherence to single-page formats. Rather than contradicting the principle of clarity, this adjustment highlights the nuanced interplay between report length and the complexity of information conveyed specific to users.

After this feedback, the most recent version (V3) was developed as shown in Figure 1. Its updates included changing the section title ‘’Call to Action’’ to ‘’Measures Under your Control’’ to improve clarity and agency. It also included separating sections (‘’Measures Under Your Control’’ and ‘’Outcomes’’) to two different pages while keeping the third page for the legends and the recommendations section. Other changes included using a single color per graph and reorganizing the “Your Complete Results” section to integrate both longitudinal actigraphy data and self-reported scores (PHQ-9 and GAD-7) over the same time period. Other minor formatting changes were also made: the legend section was moved to a third page to reduce visual density of the results section. All graphs were standardized to use the same X-axis (time), providing for an easier tracking of symptoms.

At the time of writing, our reports take the form of static PDFs. This was chosen in order to better align with current practices in our clinics, which lack a modern electronic medical record and where paper charts and paper or PDF blood test results are still common and familiar to both clinicians and patients. Based on user feedback and findings from the literature, we aim to transition to web-based formats for future iterations which would have the additive potential of being more dynamic by, for example, including real-time data streams (Chishtie et al., 2022; Turchioe et al., 2019). One positive aspect of the current focus on PDFs would be the applicability of our report format to low-resource settings.

### Early Feedback on the Report

In parallel to Design Committee feedback, semi-structured Experience Interviews were conducted with participants of the DeeP-DD study after they received a report to gather feedback. Respondents consistently described the reports as clear and helpful. One participant remarked:

> *“The report explained the data very well. I really like these things so the more data the better. But I’m not the average Joe… so I think for the average person the report has everything they would want… it is concise and has all the information a person would want.”*

Participants also reported strong trust in the information, especially when it aligned with their own experiences:

> *“It lines up with how I am feeling, so I trust it. I fully trust the sleep and activity measures because those are objective… For example, that first month I was going through a depressive cycle and wasn’t working out or socializing, and that was reflected in the report… it was a good reminder to get back.”*

User feedback also mirrored Design Committee observations, highlighting the need to reduce dense content on the first page, increase font size, and improve visual organization. Although the use of color was appreciated, limiting its variation (i.e. not having too many different colors on one page) was suggested to enhance readability.

Although the vast majority of user feedback was very positive, a critical comment emerged regarding the perceived transparency and reliability of certain data sources. As one participant noted, *“Before, when it was the watch, parts of me were like, I don’t understand how they’re getting this information… I was, like, a little bit wary,”* indicating that trust increased only when the data felt more concrete, such as *“the app tracking, the number of phone calls or screen time.”*

Feedback remained positive for the second round of reports; participants continued to find the information clear and useful. Reflecting on their clinicians’ perspectives, answering the question ‘’Did you feel your clinical team trusted the information in the report they got? Did you feel they understood it?’’, one participant emphasized the value of collaborative engagement:

> *“I think so. Yes, they seem to have a lot of confidence in it and really be motivated by it. Their motivation made me more confident and interested in the data too. Their attitude mattered more than the result itself. That was what helped the most, the attitude they had toward me based on the study and the tools they were using.”*

It is worth noting that, although the literature emphasizes the importance of remaining attentive to potential ethical concerns when returning results to patients, no such issues have emerged in the early feedback collected so far.

To complement this qualitative user feedback, a Patient Experience Questionnaire was also administered monthly via a secure REDCap link. A summary of the results from these questionnaires can be found in Table 2. The findings suggest that, in this initial piloting phase, the reports were largely regarded as helpful, simple, clear and trustworthy, but that further work is needed to increase perceptions of clinical utility. It should be noted that these scores were obtained when the clinicians and patients involved were also getting used to using the reports in practice; in addition, several of the patients involved were relatively stable. As such, with both more experience in report use and further iteration of the report, as well as use when patient symptoms are in greater flux, we may expect to see increasing perceived utility.

**Table 2.** Summary of Patient Experience Questionnaire Responses.

| Question/Item | Mean (M) | Standard Deviation (SD) | Sample Size (n) | Scale (1–5) |
| --- | --- | --- | --- | --- |
| Trust in information shared with clinical team | 4.00 | 1.32 | 9 | 1 = Not at all, 5 = Very much |
| Perceived usefulness of report | 3.25 | 0.89 | 8 | 1 = Not useful at all, 5 = Very useful |
| Ease of understanding | 3.25 | 0.89 | 8 | 1 = Not easy to understand at all, 5 = Very easy to understand |
| Representation of progress over time | 3.00 | 0.53 | 8 | 1 = Very poor representation, 5 = Very good representation |
| Amount of information provided | 2.88 | 0.64 | 8 | 1 = Far too little, 5 = Far too much (3 = Just right) |
| Clarity of summary sections | 2.88 | 0.64 | 8 | 1 = Not helpful at all, 5 = Very helpful |
| Report helped clinical team manage care | 2.56 | 1.42 | 9 | 1 = Not at all, 5 = Very much |
| Emotional distress from data access (*reverse scored*) | 4.00 | 0.87 | 9 | 1 = Very much, 5 = Not at all |
| Perceived empowerment to manage health (*reverse scored*) | 3.50 | 1.31 | 9 | 1 = Very much, 5 = Not at all |

At least two instances of clear clinical utility, from the clinician and caregiver perspective, were forthcoming. One patient with a history of psychosis was assessed by their clinician on the day they returned their actigraphy watch and, during that visit, the clinician decided the patient needed to be hospitalized because of a relapse of psychotic symptoms. After reviewing their actigraphy data with the patient and family at the next outpatient visit after the patient had stabilized, the clinician was able to identify increased steps (related to the patient pacing) and reduced sleep in the few days before the hospitalization, which served as useful markers for the family to assess when monitoring for acute changes in the patient going forward. More recently, examining the phone use data derived from the phone app with a patient with bipolar disorder revealed that the patient was using their phone primarily in the mornings; inquiring about this pattern, the clinician discovered that the patient was suffering from akathisia when they woke up, and was coping using their phone to distract themselves. This led to improved treatment of akathisia and discussion of side effects and medication choices.

A fundamental component of the study is to actively involve patients in their own care by providing them access to their personal data, an approach that, unfortunately, is rarely implemented in most research projects (Long et al., 2016). The delivery of these reports was intended to enhance patients’ understanding of their clinical states, serving either as a reinforcing tool when outcomes were positive or as a mechanism to help identify behaviors that could be detrimental to their health trajectory. An anecdotal example this happened with Patient 014, who had been diagnosed with non-psychotic bipolar disorder approximately a decade prior to enrollment, already possessed considerable awareness of factors conducive to clinical stability, as well as those that posed risks. She recognized, in particular, the importance of adequate sleep. However, upon reviewing her actigraphy data, she discovered that her average sleep duration was limited to approximately five hours per night. This objective feedback prompted her to establish a personal goal of improving her sleep and adopting more consistent sleep practices.

## Discussion

This study presents an initial effort to address a critical yet underexplored aspect of digital phenotyping in psychiatry: the effective communication of collected data back to patients, clinicians, and caregivers. Our findings further add to the literature underscoring that digital phenotyping technologies can indeed support clinical care, patient engagement, and collaborative management when data are thoughtfully returned using evidence-based design principles.

The results from our narrative literature review align with growing evidence that patients value access to their personal health data when presented clearly and intuitively (Krägeloh et al., 2015; Sayeed et al., 2021). Indeed, structured and visually appealing data reports, such as those developed in our iterative design process, were positively received by patients, enhancing their sense of agency and promoting active engagement in care. This mirrors broader literature suggesting that integrating patient-reported outcomes with passively collected digital biomarkers can enhance patient-centered care and foster therapeutic alliance ((Chang et al., 2023); (Carlier et al., 2012; Insel, 2017)). Consistent with previous findings (Krägeloh et al., 2015; Sayeed et al., 2021); (Polhemus et al., 2022)), our results emphasize the need for individualized, flexible reporting options that allow patients to provide feedback on the depth and detail of data they receive.

Our results also highlighted important considerations already discussed in some of the literature (Sayeed et al., 2021): the complexity and volume of data presented must be balanced in order to avoid reduced understanding and potential emotional distress, particularly for sensitive or ambiguous data. To date, user feedback suggests that the reports achieve an effective balance, with comments indicating minimal distress and an appropriate amount of information. However it is worth mentioning that, although still in the early stages of report development, some feedback indicated that more work is needed to enhance patient empowerment and perceived clinical utility. It is likely that this is partially due to this being the early phases of these reports being integrated into clinical care as well as the lack of all possible measures (e.g. the app-related measures). Interestingly, some feedback from the Design Committee diverged from the literature by mentioning that the use of numerous colors and a dense layout could limit comprehension. As part of our iterative approach, certain modifications were therefore necessary, even if they were different from established practices in the literature, such as extending the report across multiple pages to improve clarity and user understanding.

Our iterative development process, informed by (Midway, 2020)’s visualization principles, underscored the value of simplified, focused formats, such as line graphs and traffic-light color schemes, in promoting intuitive communication. Preferences for these elements emerged consistently in patient and committee feedback and align with broader evidence in healthcare visualization literature (Hartzler et al., 2015; Lor et al., 2019; Turchioe et al., 2019), reinforcing that visual design serves not only an aesthetic function but also significantly enhances clarity, interpretability, and engagement. In parallel, our findings highlight the ethical considerations involved in returning phenotyping results, particularly sensitive psychiatric data. Guided by (Shen et al., 2024), we adopted a tiered disclosure framework, prioritizing low-risk, actionable insights (e.g., sleep, activity, mood trends) while deferring more complex indicators until their interpretability and clinical utility are better established. This cautious approach aligns with broader ethical discourse in digital phenotyping ((Martinez-Martin et al., 2021), emphasizing participant autonomy, informed choice, and the need to minimize potential harm through unclear or premature data presentation.

When considering the return of results to patients within mental health settings, insights from authors such as (Chang et al., 2023) and (Polhemus et al., 2022) have been instrumental in guiding early efforts. Their work demonstrates that many principles established in the general medical literature, particularly those related to clarity, engagement, and perceived usefulness are also highly relevant to psychiatric populations. Consistent with these findings, our own user feedback indicates that data visualizations were viewed as central to the experience of receiving results, influencing both satisfaction and engagement, and were frequently described as helpful, simple, clear, and trustworthy. These parallels suggest that core visual design strategies, such as prioritizing clarity and intuitive formats, translate well across both general and mental health contexts. Nonetheless, the literature in this area remains limited, and we hope that our ongoing development efforts will contribute to this growing field by incorporating transdiagnostic patient perspectives and extending these insights to include physicians and caregivers.

### Limitations

Despite existing studies, the evidence supporting return-of-results design in the literature remains limited. Many studies reviewed were descriptive, cross-sectional, or qualitative, such as focus groups and interviews (Chang et al., 2023; Lor et al., 2019; Sayeed et al., 2021), rather than randomized controlled trials. As a result, while themes like patient engagement, visual preference, and data comprehension are well supported, there is less empirical evidence linking report exposure to concrete clinical outcomes. Moreover, despite strides from authors such as (Chang et al., 2023; Polhemus et al., 2022; Turchioe et al., 2019), most of the literature comes from general medical populations, not psychiatric samples, limiting generalizability. Even within studies that include psychiatric populations, such as those by (Chang et al., 2023), samples are often small and narrowly focused on few diagnoses, further constraining broader application. Further controlled studies are needed to rigorously evaluate the impact of report-based interventions in mental health care.

Our study also faced limitations. Given the preliminary and iterative nature of our report design, findings are constrained by the small sample size and qualitative and descriptive nature of feedback. Future planned research will incorporate larger-scale evaluations and randomized controlled trials to assess the clinical impact, user experience, and health outcomes associated with these personalized reports.

### Future Steps

Some next steps are worth considering. First, transitioning from static PDFs reports to dynamic, web-based interactive platforms represents a logical evolution that aligns with emerging trends in personalized mental health care, including the use of digital twins and precision medicine frameworks (M. Spitzer et al., 2023). This step is planned in the future as the overarching DeeP-DD project develops. We hope real-time feedback mechanisms, enabled by more use of mobile phone and smart actigraphy watch technologies, may offer enhanced opportunities for timely clinical intervention and personalized patient support, potentially transforming psychiatric care delivery. Also, we must highlight that the co-creation of Patient Progress Reports, through the Design Committee, Patient Experience Interviews, and Questionnaires, is still an ongoing process. While preliminary findings are promising, future versions of the reports will continue to be refined based on cumulative feedback, with the goal of tailoring content to participants’ needs. Moreover, given that most clinicians’ perspectives in the literature echoed patient feedback, particularly regarding visual clarity, data trend representation, while adding insights into the value of concise graphical summaries for rapid clinical assessment (Abudiyab & Alanazi, 2022; Khairat et al., 2018), it will be valuable to compare how their version of the report evolves and the feedback it generates as we move toward developing physician-facing formats in the next phase of the project. Finally, despite their central role in supporting mental health treatment, there is little published guidance on how to design reports for caregivers that are respectful of patient autonomy while still being useful. This notable underrepresentation in our review indicates a critical knowledge gap and an important avenue for future research (Dalstrom et al., 2025; Oakley-Girvan et al., 2023).

## Conclusion

As part of the DeeP-DD project, this review and early report development contribute to the growing digital phenotyping literature by demonstrating how evidence-based design, ethical frameworks, and user feedback can yield clear, engaging, and clinically relevant patient-facing reports. Initial versions center on patients, with future iterations planned to better support clinicians and include caregivers. Early feedback is promising, with next steps focused on broader rollout, automation, and real-time integration.

## Data Availability

All data produced in the present study are available upon reasonable request to the authors

## Author’s contribution

M.S. conceived the study and led the project design. M.S. ; MC.M ; T.Z. contributed to literature review and manuscript drafting. D.P. managed the data infrastructure for the project. D.H. wrote the scripts used to generate graphs presented in the reports. T.Z. ; I.G. ; D.H. participated in data extraction. C.C. ; K.L. ; D.RC. ; T.W. ; L.P. ; B.S ; D.P; S.J. ; G.S ; T.Z. ; MC.M. and S.K helped review the manuscript. D.B. reviewed the manuscript, provided supervision, and helped conceive the study. All authors reviewed and approved the final manuscript.

## Acknowledgements

We thank the members of the lived experience councils of the Center of Excellence in Youth Mental Health at the Douglas Mental Health University Institute for their support and feedback throughout this project. We are also grateful to participants and the Design Committee* for their valuable contributions.

Design Committee Members*: Dr. Delphine Racher-Chéné; Dr. Jai Shah; Dr. Katie Lavigne; Dr. Anthony Gifuni; Dr. Vincent Paquin; Dr. Soham Rej; Dr. Stefan Kloiber; Astrid Alice Bastin; Raven Erin; Béatrice Schunn and other members of the Design Committee

## Supplementary Material

**Figure 1.**
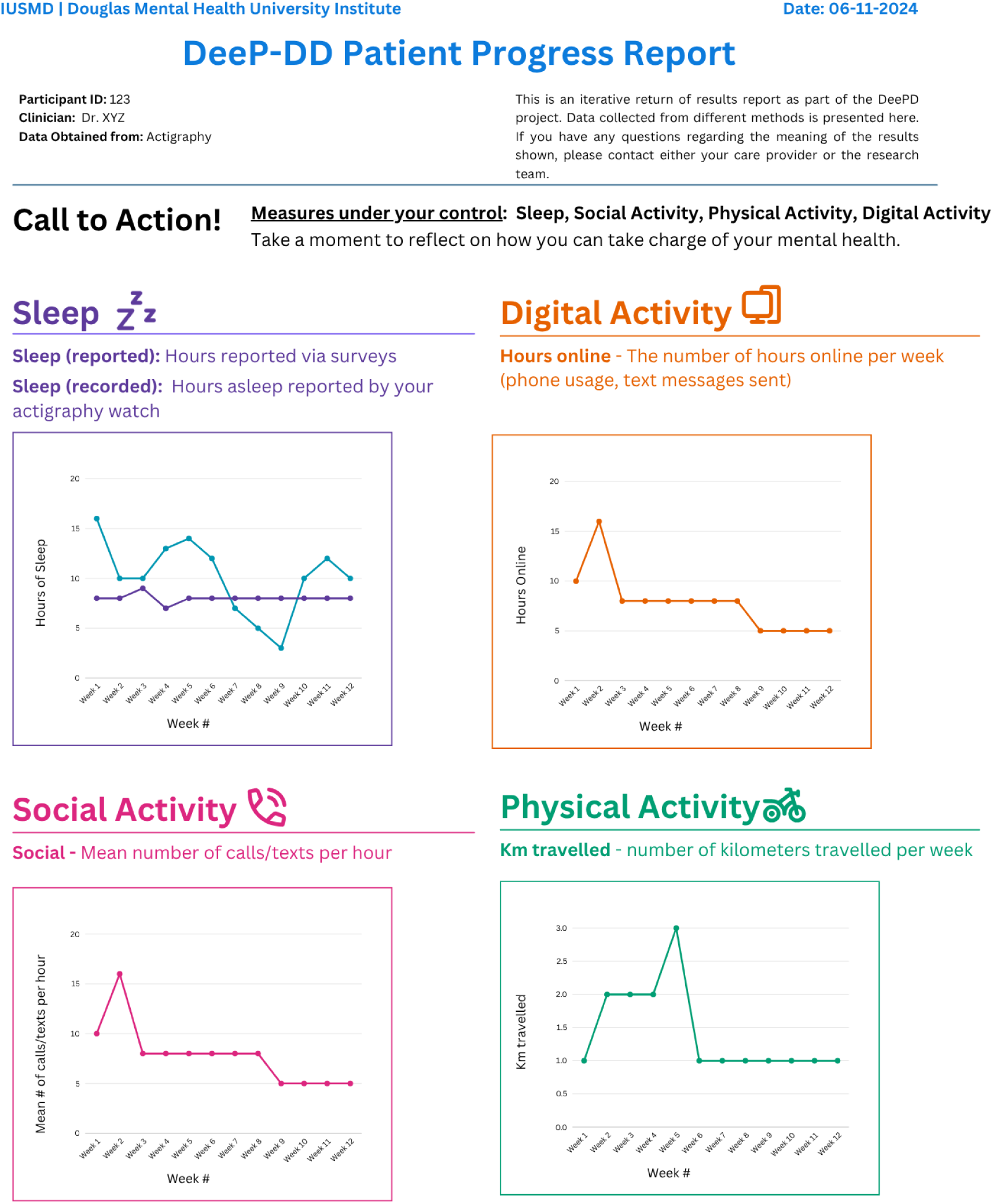
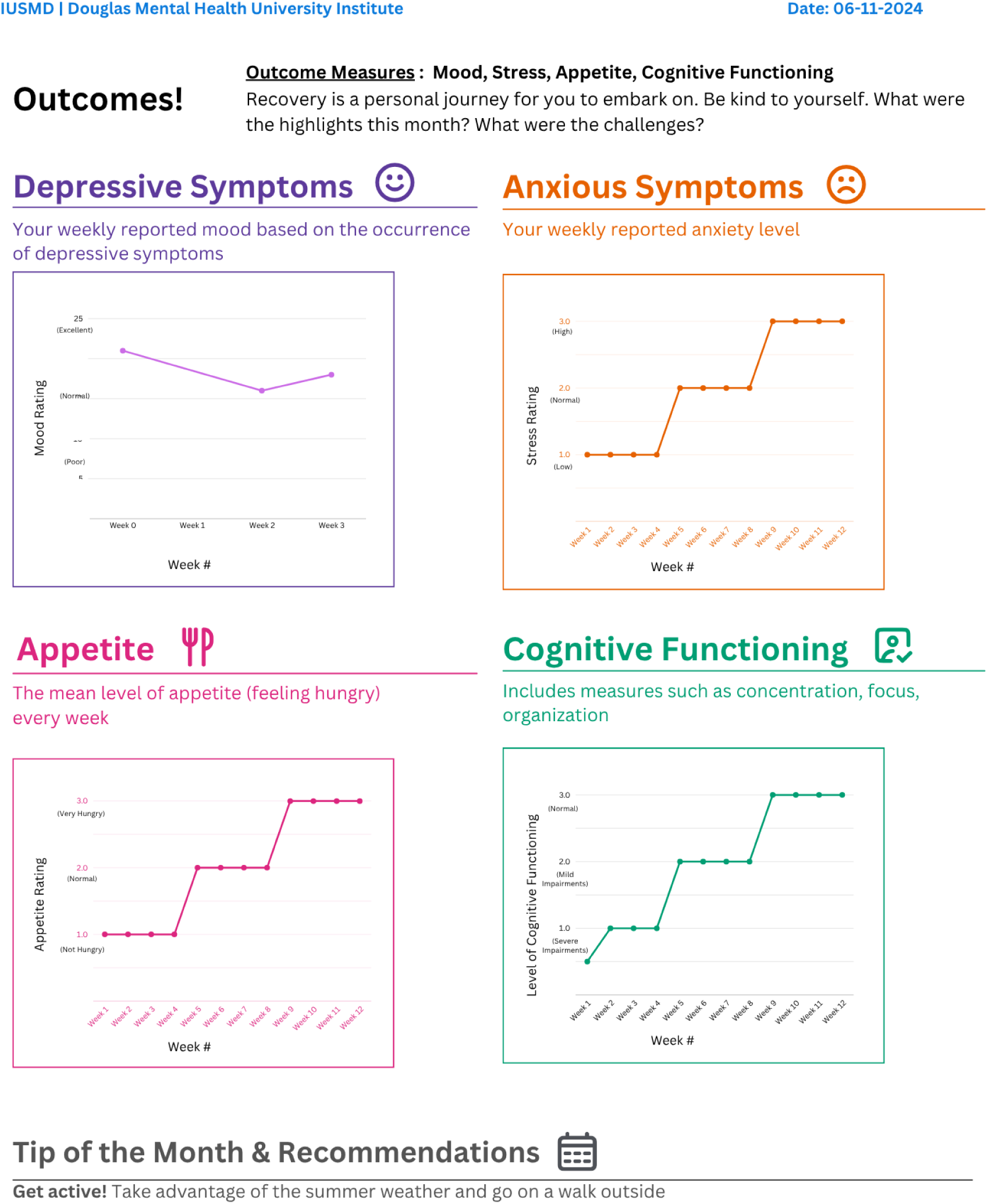
Mock version (June 2024)

**Figure 2.**
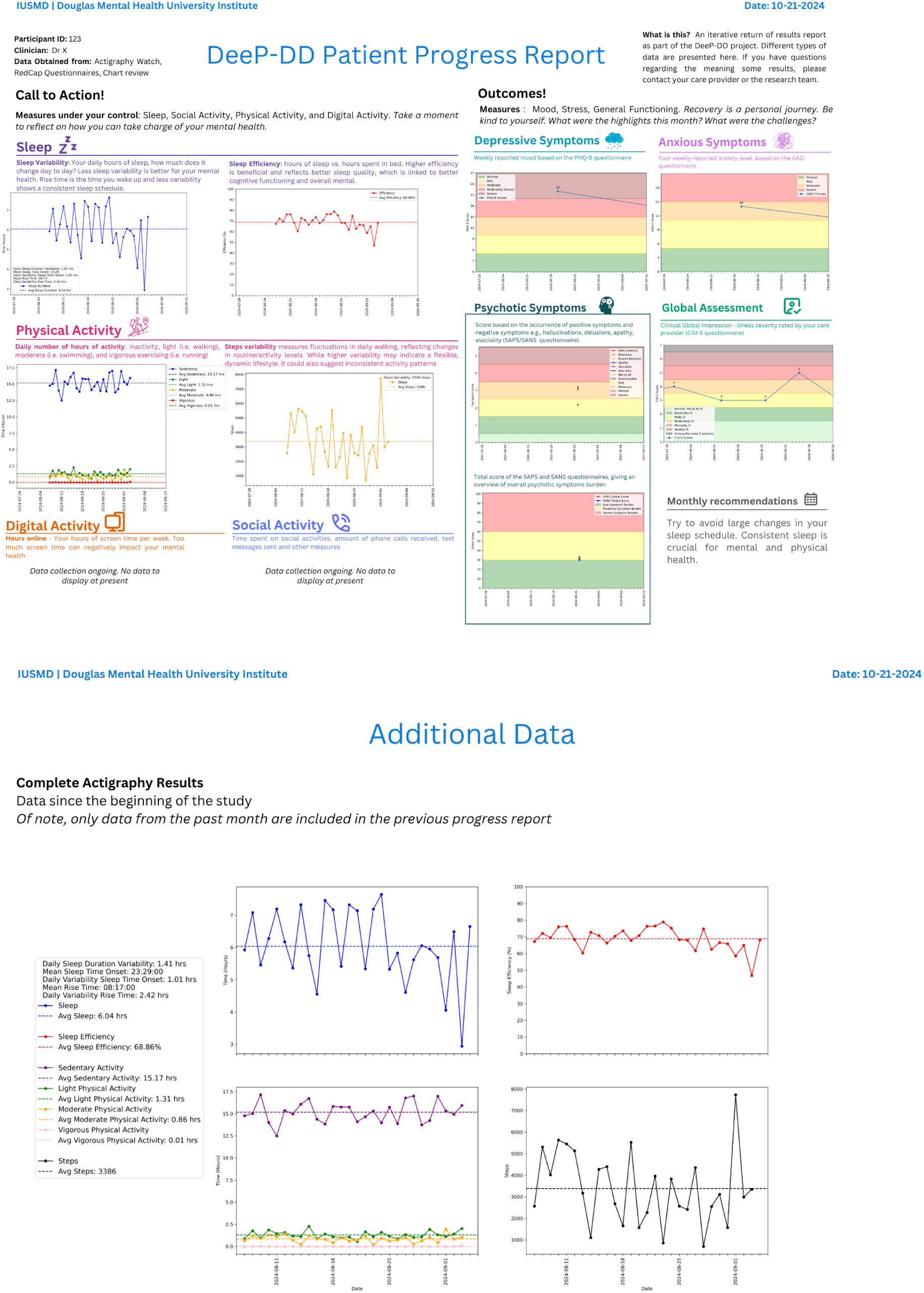
Patient Report Version 1. **Version 1 Report Description:** Divided into three sections on a double-sided page. Right side of front page: section (1) “Call to Action” with monthly actigraphy measures from worn trackers, modifiable by patient (e.g., sleep hours). Left side: section (2) “Outcomes”: data from self-reporting and clinician-rated scales (e.g., PHQ-9 score for depressive symptoms) with traffic light color-coded backgrounds reflecting score severity. Each category of data in (1) and (2) included an icon and a short label written in a distinct color for clear demarcation. Bottom right of front page: monthly recommendation based on (1). Back page: section (3) “Additional Data”: actigraphy trends since study start. All data in Version 1 were presented as line graphs.

**Figure 3.**
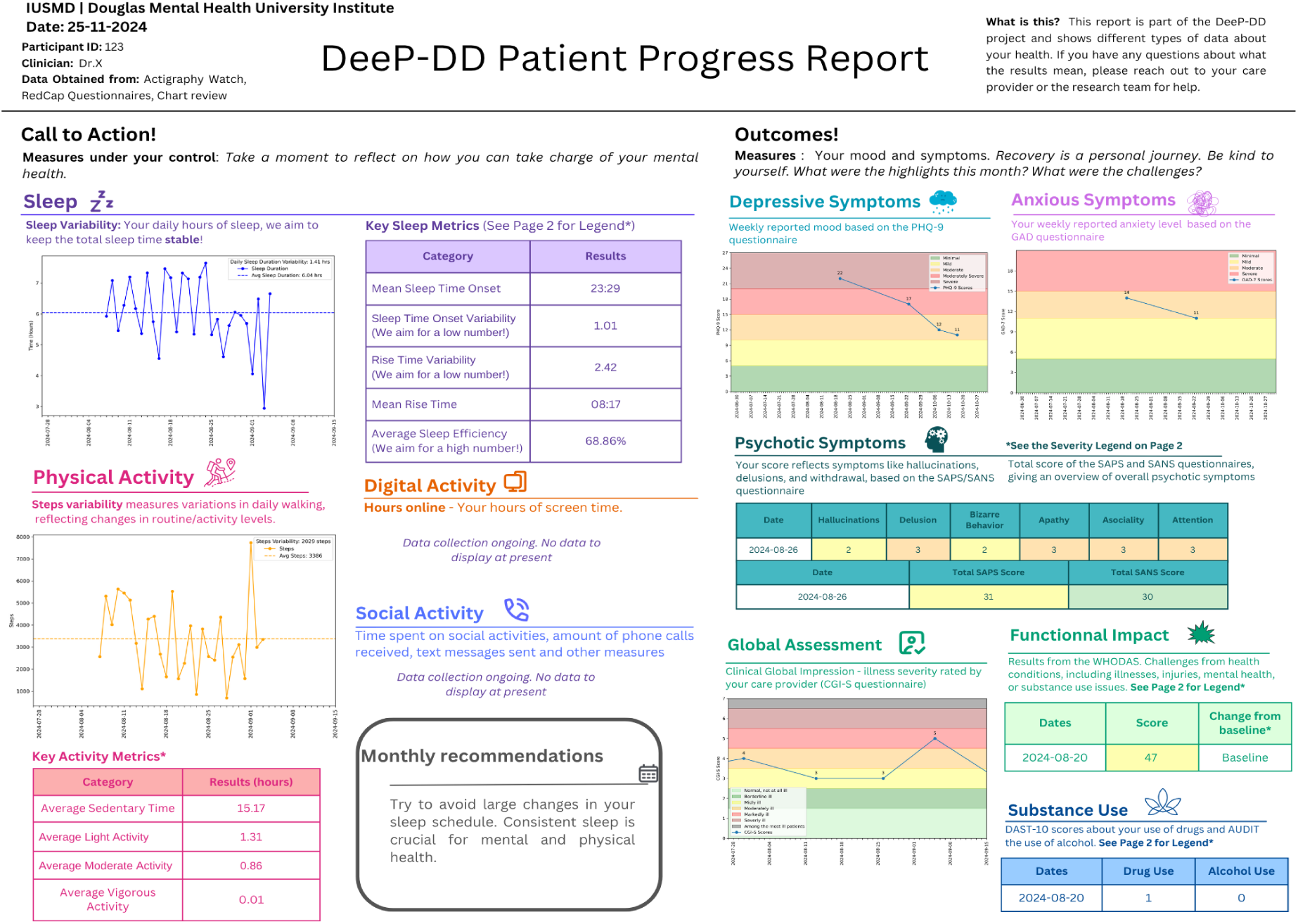
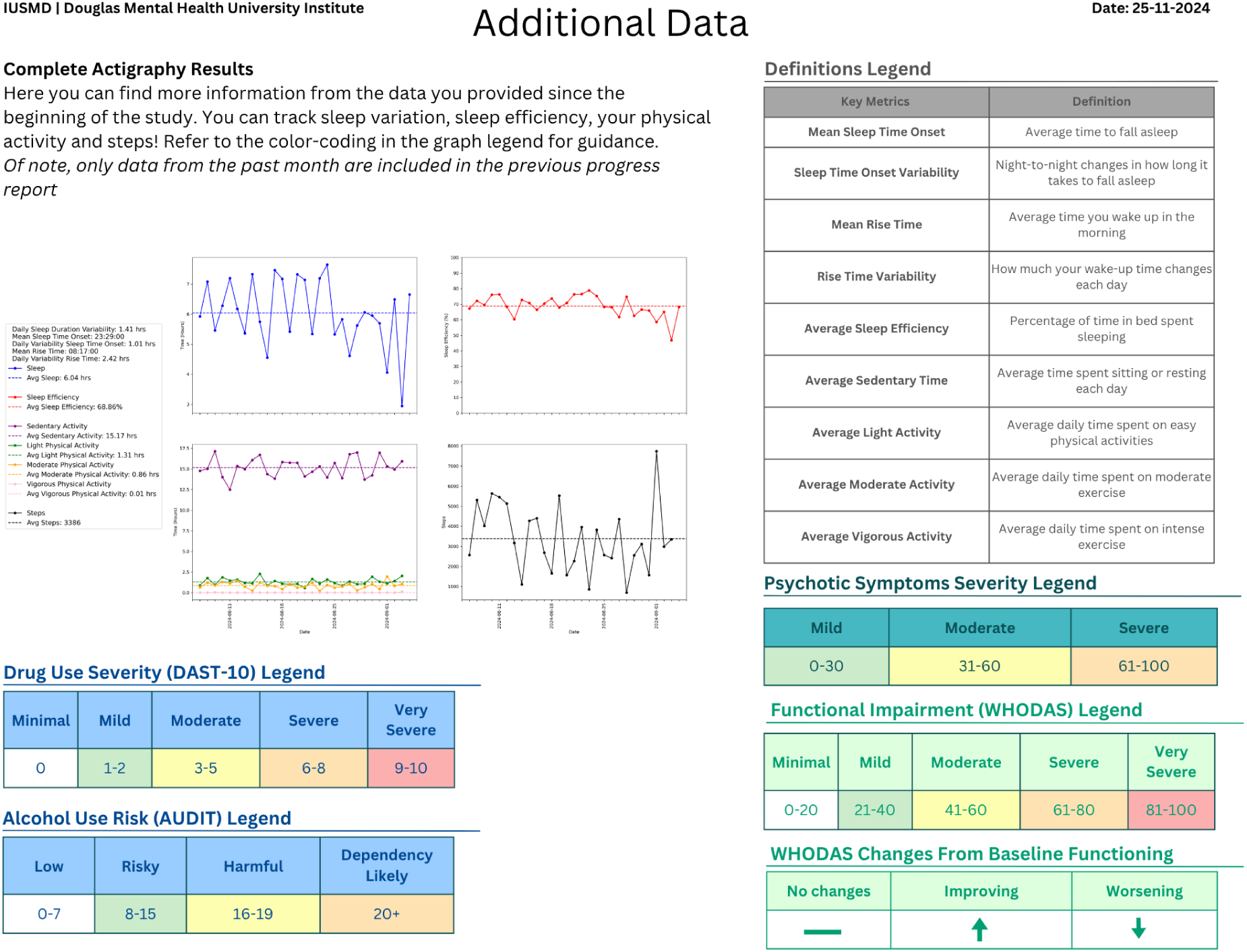
Patient Report Version 2. **Version 2 Report Description**: Sections division similarly to V1. Back page “Additional Data” section modified to include severity scale legends and definitions of key metrics. Some categories of data in (1) (e.g., sleep efficiency) converted from line graphs to tables and additional data included in tables (e.g., mean rise time). In (2), some graphs are also converted to tables, while keeping with the traffic light color scheme to help track symptom severity.

**Figure 4.**
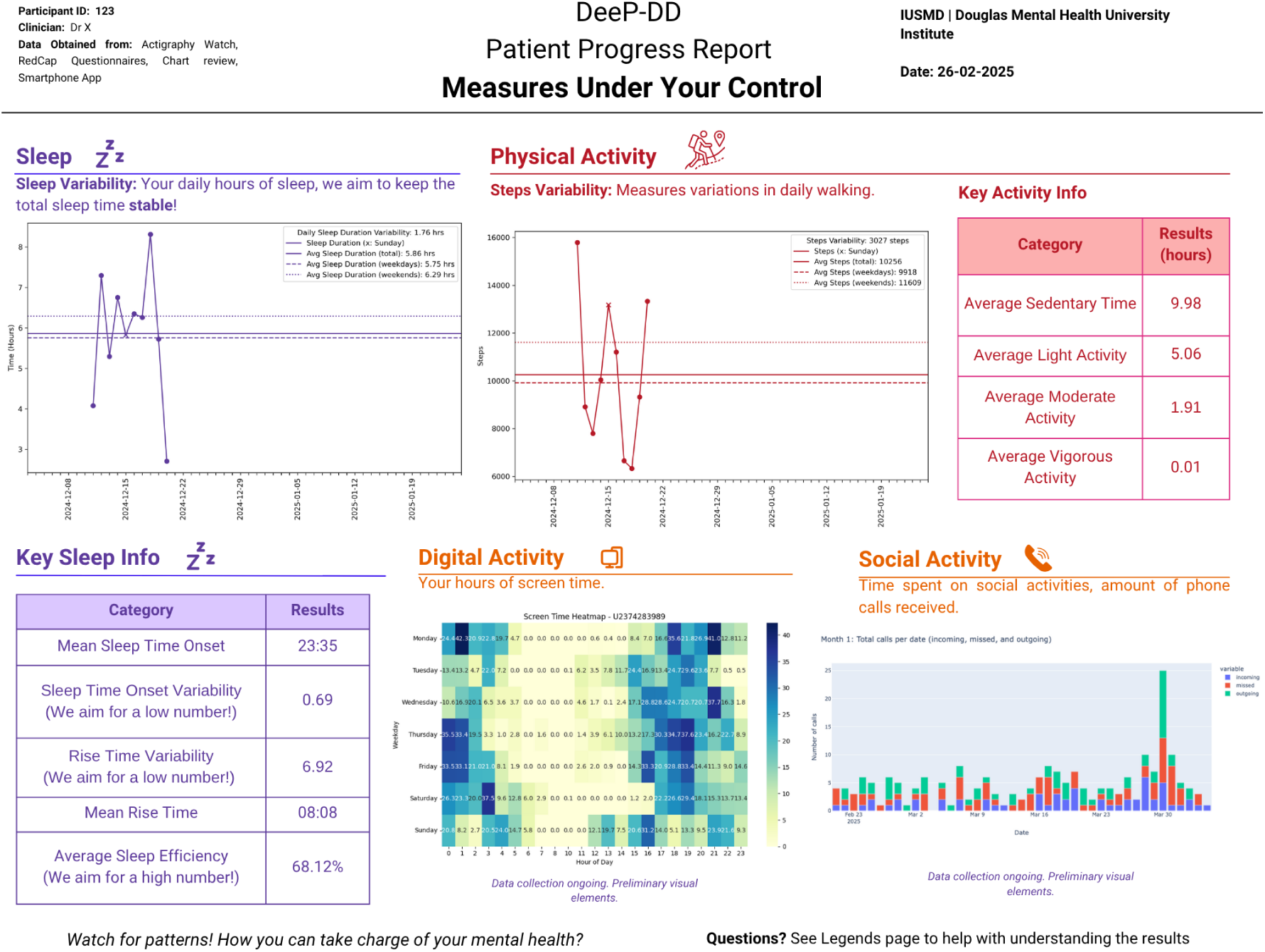
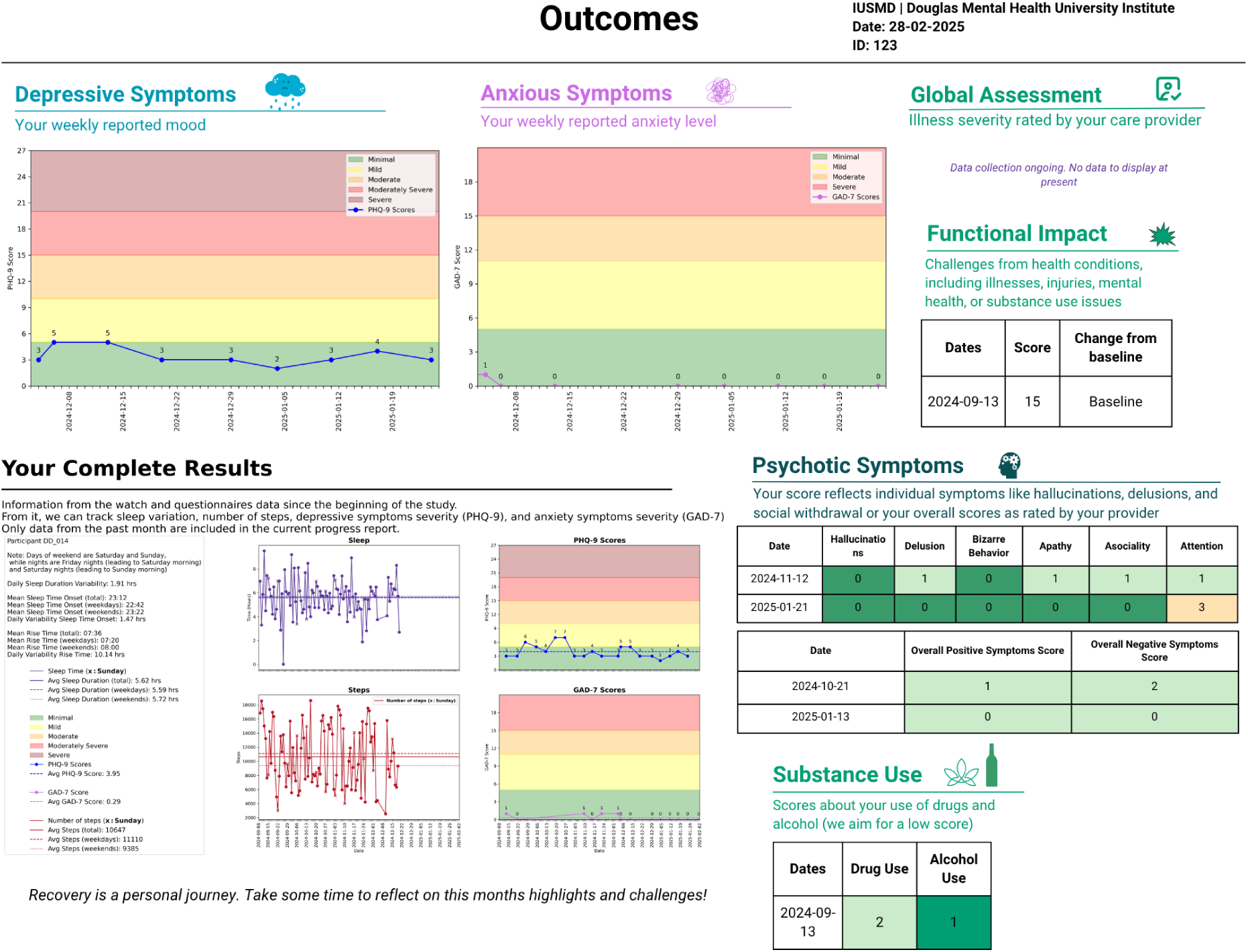
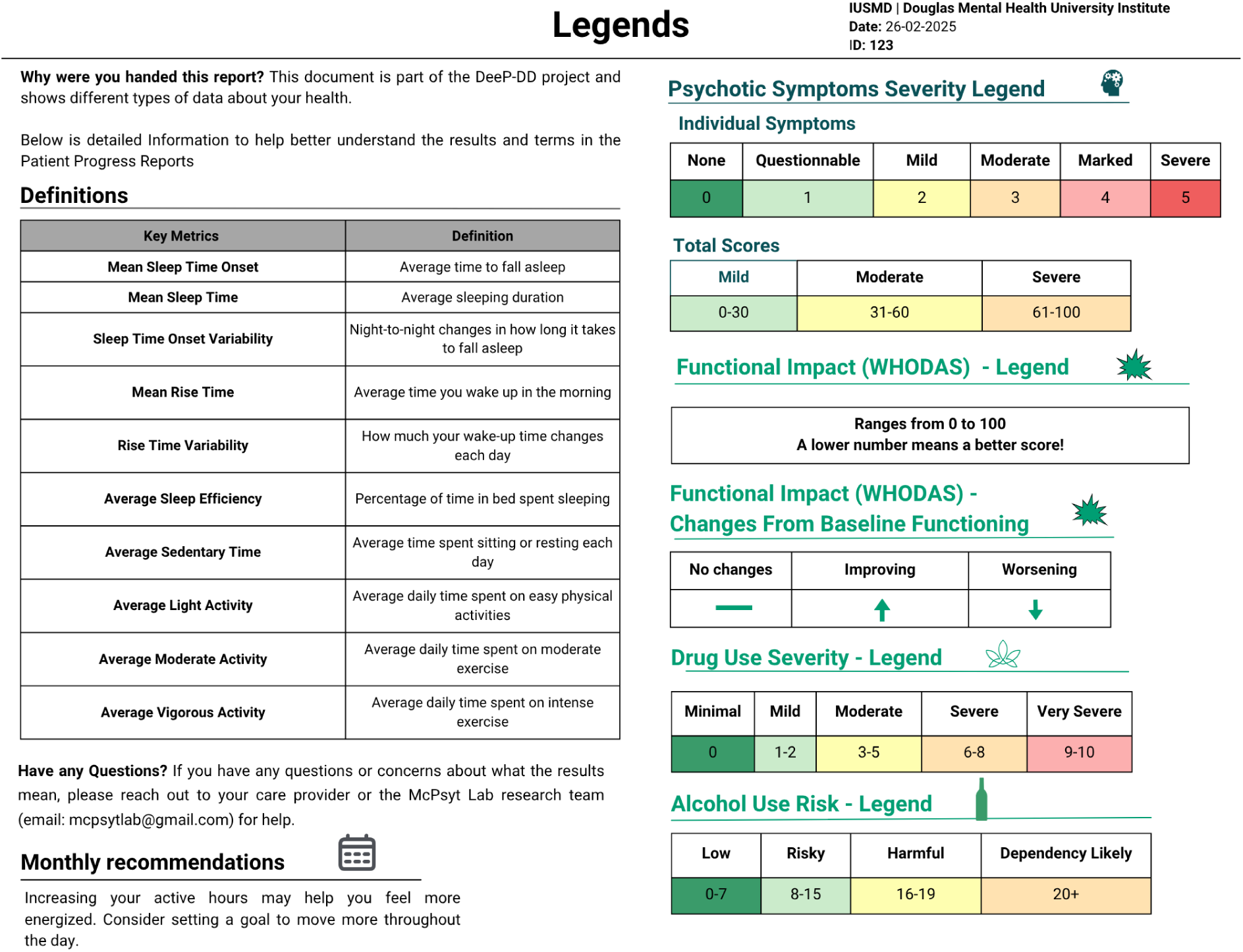
Patient Report Version 3. **Version 3 Report Description**: Report content division revised and distributed across three single-sided pages to improve readability. First page: (1) “Call to Action” renamed “Measures under your control”. Preliminary visual elements for “Digital” and “Social” Activities were available but not finalized at the time of writing. Second page: (2) “Outcomes” and (3) “Additional data”, renamed “Your complete results” – bottom left – and now comprising self-reported scores alongside actigraphy data from study beginning. Sundays are marked with an “x” on line graphs to highlight differences between weekends and weekdays. Graphs display mean values for weekdays, weekends, and overall totals, allowing clearer interpretation of patterns across time. Both self-reported/clinician-rated score data tables and line graph keeping with the traffic light color scheme. Third page: additional section (4) “Legends” with severity scale legends and definitions of key metrics. Monthly recommendation section also moved to the last page bottom left. Each category of data still includes an icon and a short label written in a distinct color. Report’s colors reviewed to ensure consistency between each symptom category, particularly in the “Your Complete Results” section. All graphs standardized to use the same X-axis.

**Figure 5.**
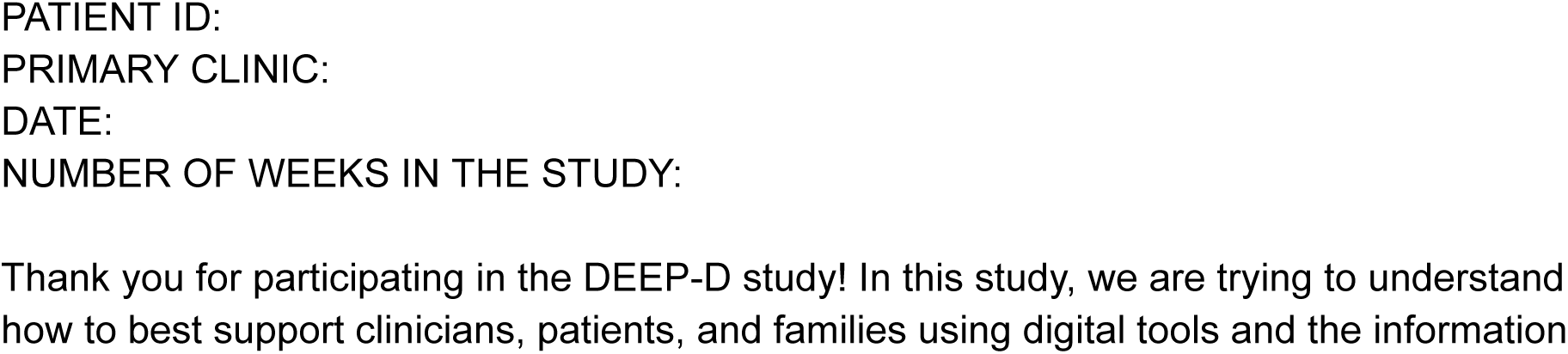
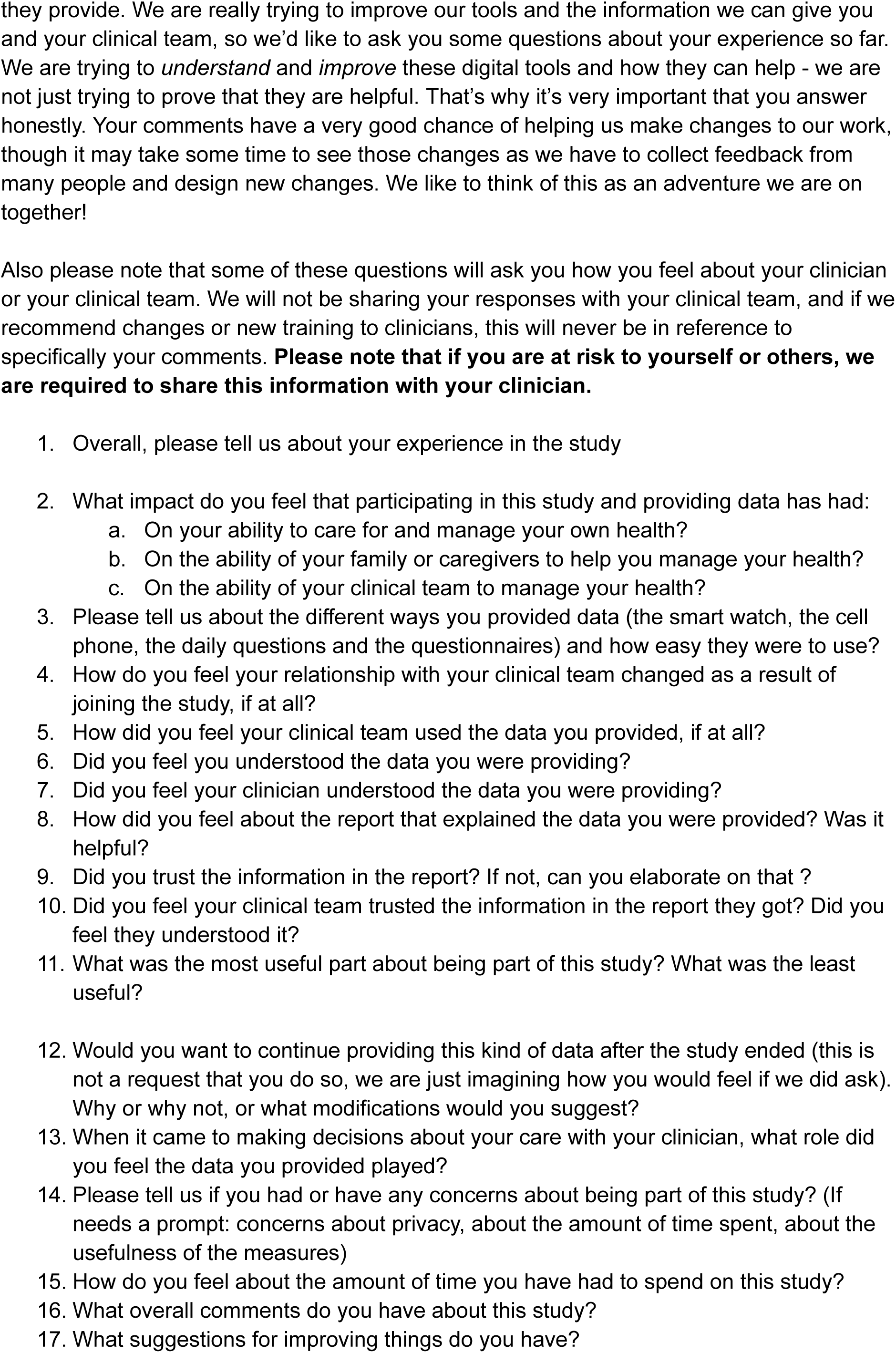
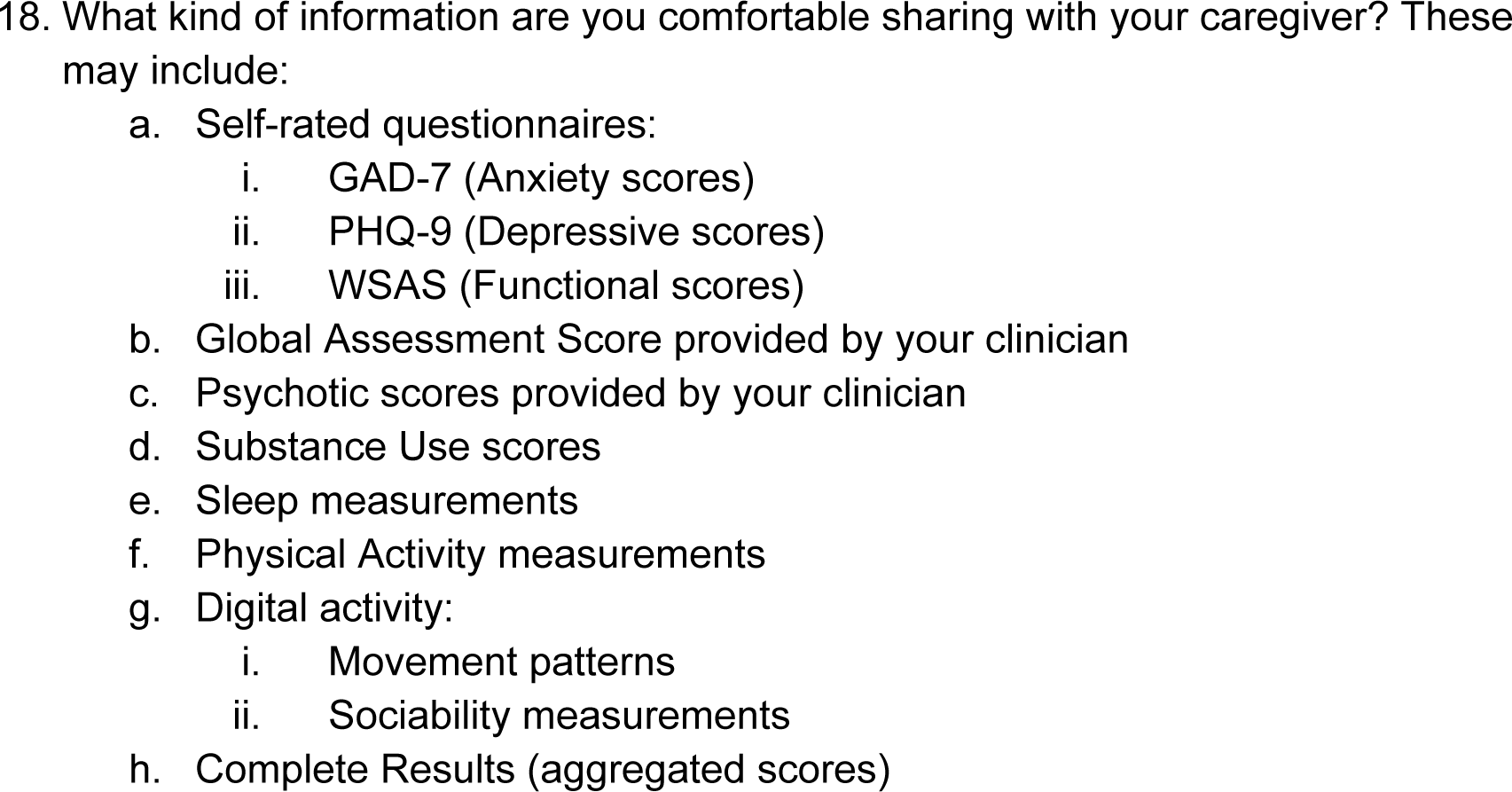
Patient Experience Interview.

**Figure 6.**
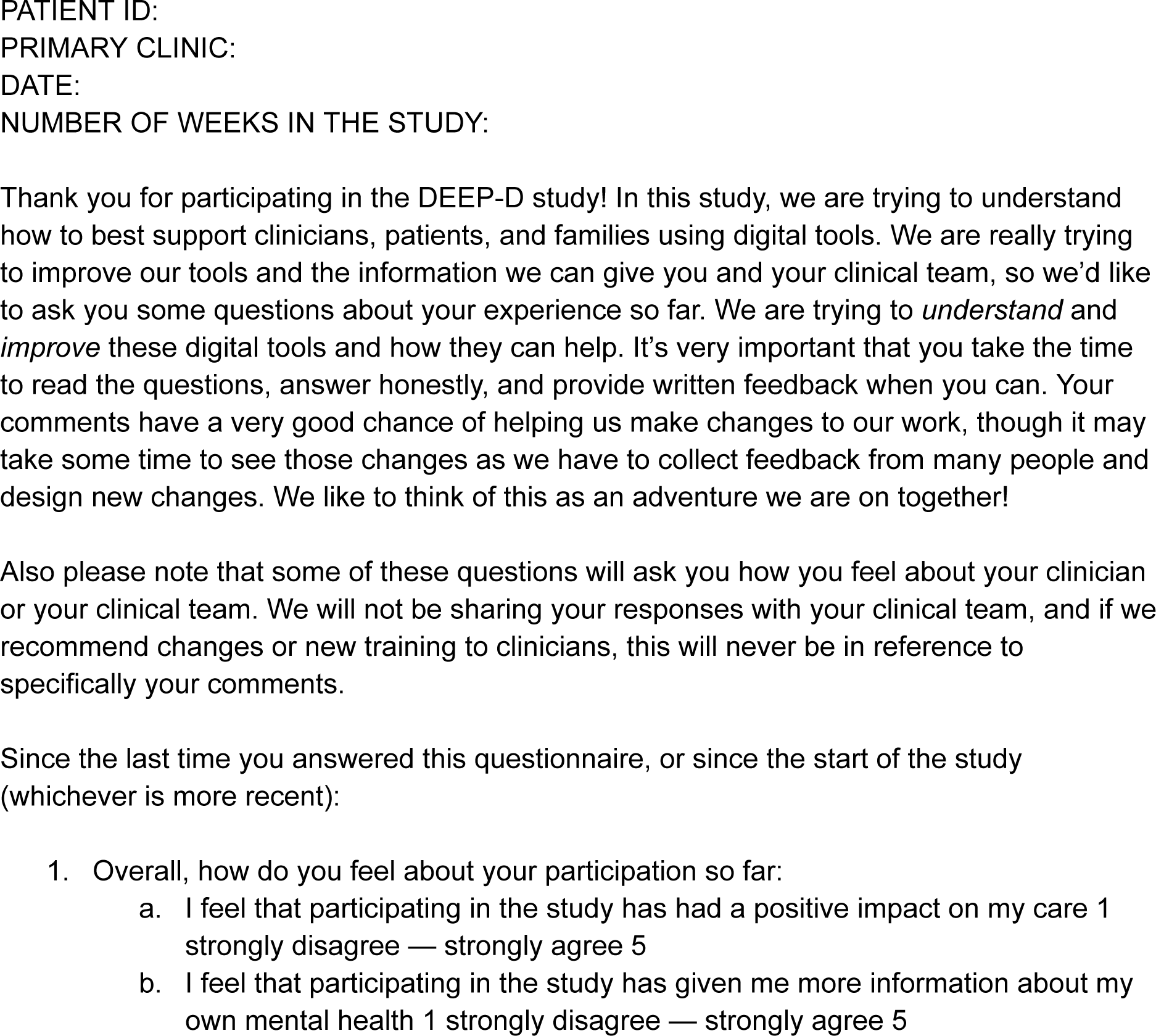
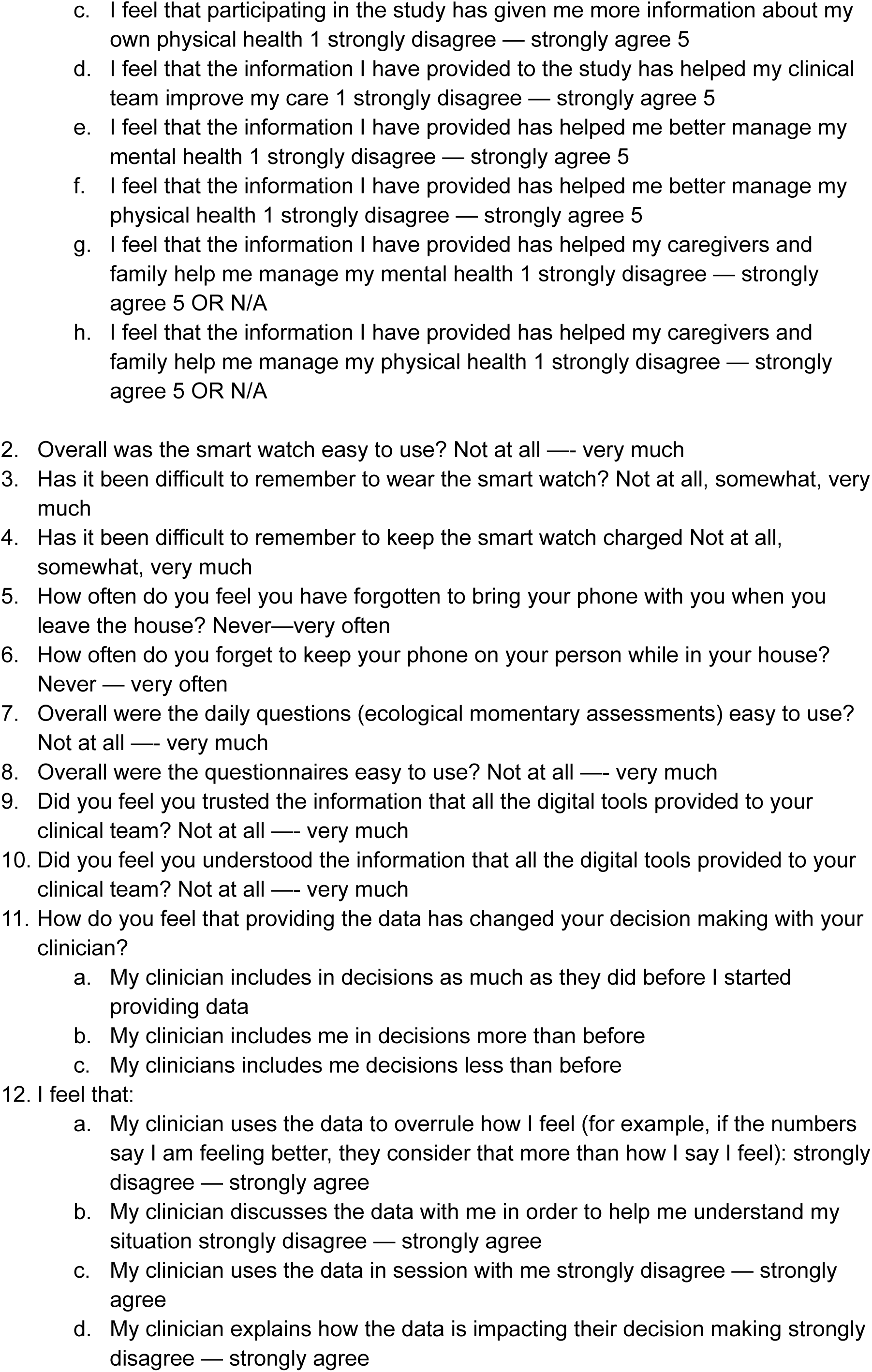
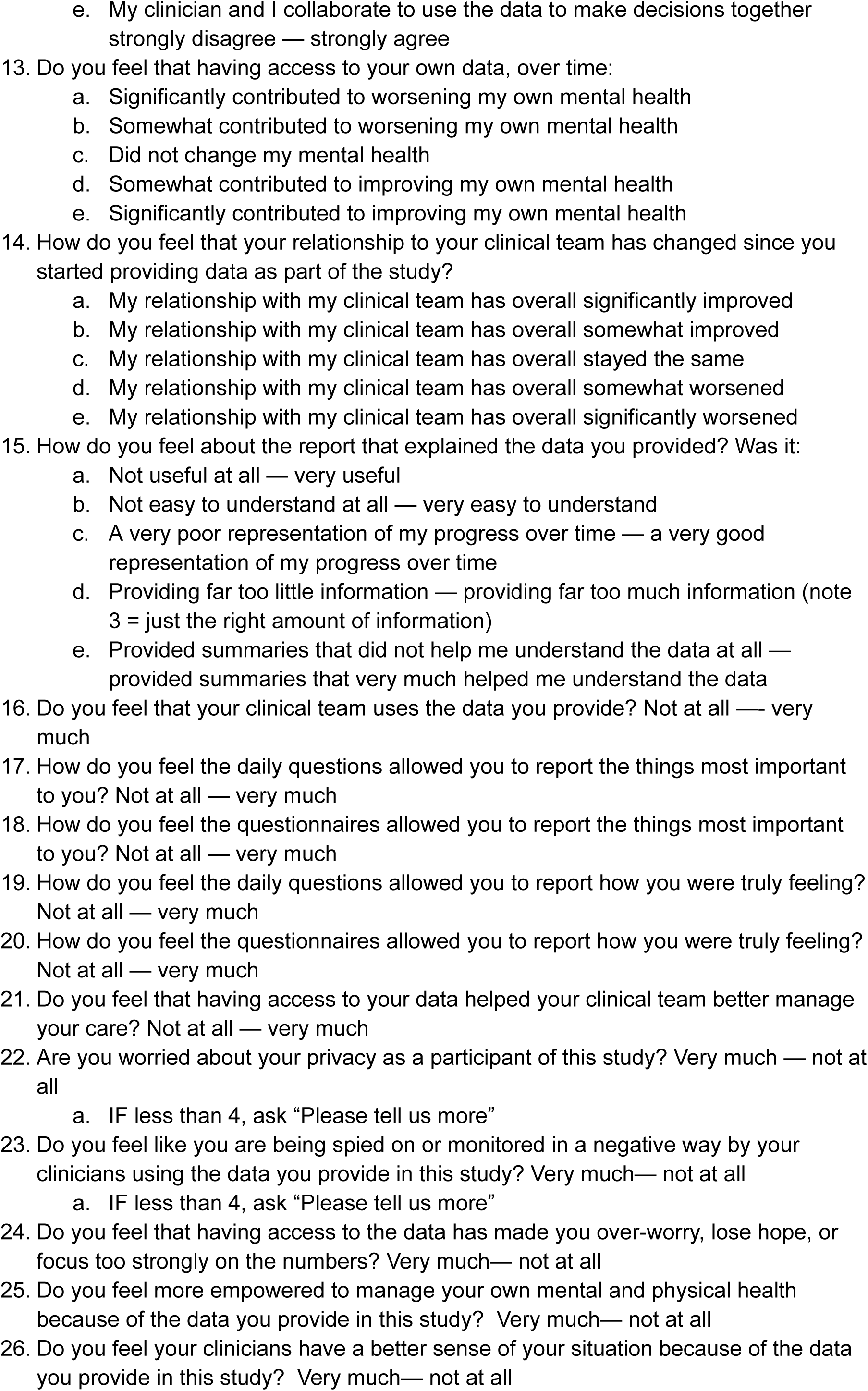
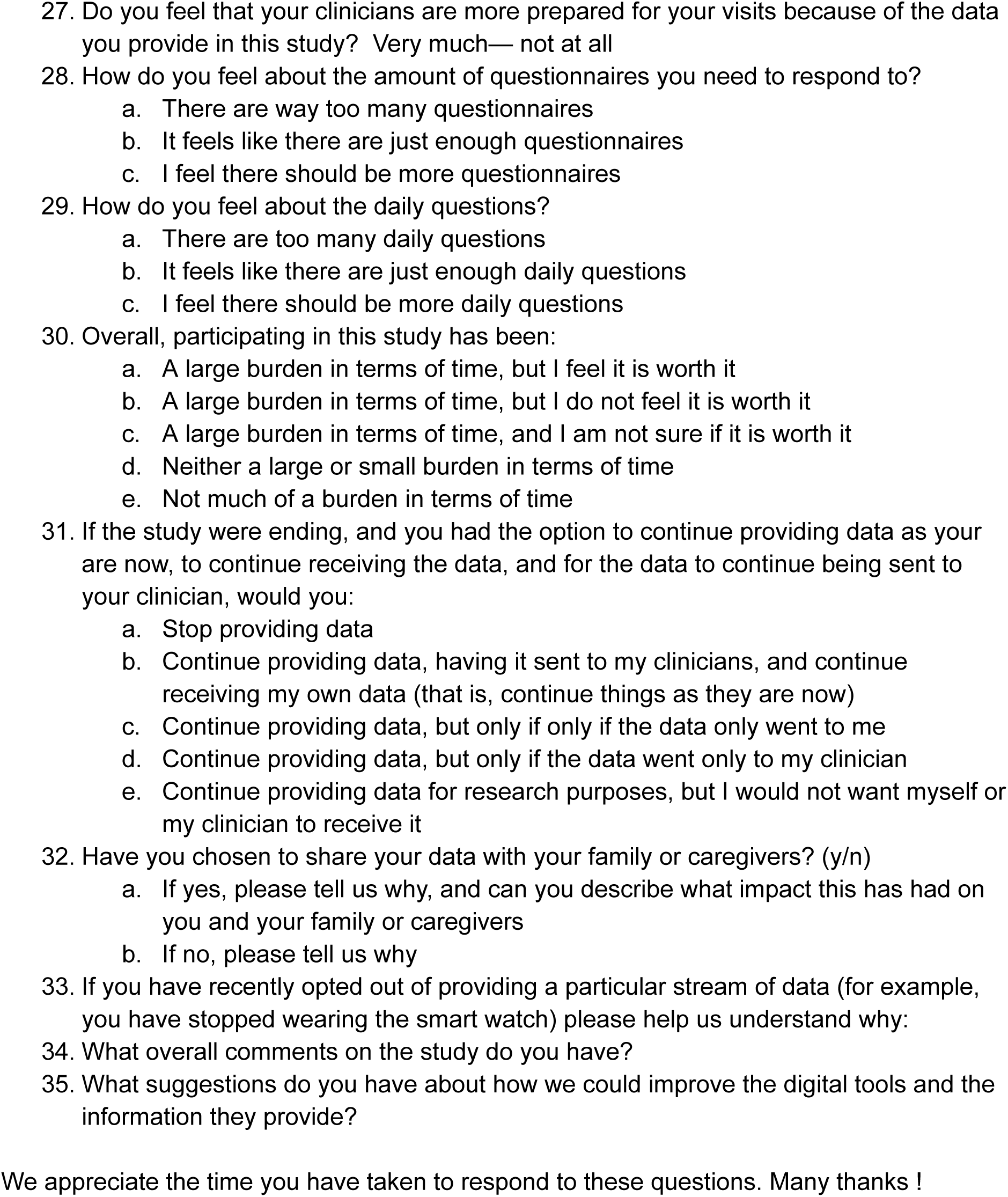
Patient Experience Questionnaire.

